# Infant Mental Health Prevention and Treatment: A Systematic Review Examining the Role of Homelessness

**DOI:** 10.1101/2020.12.09.20246553

**Authors:** Taylor D. Landis, Megan M. Hare, Paulo A. Graziano

**Affiliations:** Center for Children and Families, Department of Psychology, Florida International University

**Keywords:** infancy, mental health, intervention, homeless

## Abstract

**Purpose:** Whereas many prevention and treatment programs exist for children and families, there have been no reviews specifically examining infant mental health outcomes. Furthermore, despite high rates of infants and families experiencing homelessness, little work has evaluated the effectiveness of these parenting programs in such vulnerable populations. Therefore, the purpose of this study was to a) systematically examine prevention and treatment parenting programs targeting infant mental health outcomes in infants from birth to age 2 years, b) highlight strengths and limitations of current interventions, and c) identify gaps in the existing literature to inform future mental health intervention science in areas of greatest need, specifically within the context of homelessness.

**Methods:** From over 15,000 publications initially identified, thirty-three prevention and treatment programs met inclusion criteria for this review. Each program was reviewed for level of scientific evidence.

**Results:** Of the thirty-three programs reviewed, eleven (33%) were classified as promising. An additional 18% were classified as ineffective, emerging, and effective. Lastly, only four programs (12%; Attachment and Biobehavioral Catch-Up, Parent-Child Interaction Therapy, Triple P-Positive Parenting Program, and Video-feedback Intervention Parenting Program) were classified as evidence-based based on infant mental health outcomes. Few of the identified programs have been implemented in homeless shelters, with no randomized control trials to date.

**Conclusions:** There is a dearth of literature examining programs targeting infant mental health. Even those programs considered evidence-based have not been thoroughly examined among families in shelter settings.

---

Over half a million people in the United States are homeless each day, with approximately 35% of those individuals experiencing homelessness unsheltered (www.whitehouse.gov/cea). The prevalence of homelessness has increased in recent years, including greater numbers of children (Bassuk, Richard, & Tsertsvadze, 2015; Homelessness, 2020). A resounding 2.5 million children, or 1 in 30, in the United States experience homelessness every year (https://www.air.org/center/national-center-family-homelessness). Mental health problems occur at higher rates among children and families experiencing chronic homelessness (Lippert & Lee, 2015). Specifically, mothers who have recently become homeless report having greater rates of mental health problems, especially symptoms of trauma (Bassuk, Buckner, Perloff, & Bassuk, 1998; Zima, Wells, Benjamin, & Duan, 1996). Furthermore, mothers report having difficulty coping with homelessness and report greater levels of hopelessness, especially regarding resources and services available to meet their needs (Tischler, Rademeyer, & Vostanis, 2007).

In terms of the impact of homelessness on child mental health, an approximated 10% to 26% of homeless preschool-aged children experience clinical levels of mental health problems, as demonstrated by a systematic review (Bassuk et al., 2015). More specifically, homeless children ages 0-5 experience more internalizing behavior problems, externalizing behavior problems, and overall mental health problems (Park, Fertig, & Allison, 2011). Unfortunately, despite the dire need for prevention, intervention, and housing reform, very few programs specifically target mental health among homeless populations. Given the astounding number of children experiencing homelessness, and its associated impact on child mental health, its critically important examine programs designed to prevent and treat mental health problems in young children who are at-risk.

Consistent with a systematic review by Bagner and colleagues (2012), infancy is defined as children under 2 years of age. The first two years of life are considered a sensitive and critical developmental period, in which neural plasticity occurs at greater rates establishing important circuitry architecture in the brain (Uylings, 2006). During this critical period, children are more susceptible to robust neurological impacts from environmental factors. Relatedly, environmental stressors within the first few years of life can have longstanding deleterious impacts on children’s development, including cognitive functioning, behavior, language, and social-emotional development (Knudsen, 2004; Uylings, 2006). Homelessness certainly qualifies as a potentially high environmental stressor for families, which may have direct and indirect consequences to children’s development, particularly during infancy. Therefore, maximizing environmental support to optimize development during the first two years of life and/or buffer some of the negative effects of a high environmental stressors, is of utmost importance.

## Mental Health Needs of Infants

The most common problems during infancy include emotional, behavioral, sleep, and eating difficulties, along with child abuse and neglect (Carr, 2019; Skovgaard et al., 2007). Importantly, there are many challenges and limitations to identifying positive social-emotional development during infancy (Bagner, Rodríguez, Blake, Linares, & Carter, 2012; Carter, Briggs-Gowan, & Davis, 2004). Specifically, disentangling typical versus atypical behavior early in life remains difficult, with identifying and differentiating between mental health disorders adding significant complexity (Bagner et al., 2012; Foreman, 2015; Szaniecki & Barnes, 2016). Some research suggests behavior problems are normative during infancy and tend to decrease into the preschool years (ages 2-5) (Van Zeijl et al., 2006). However, other literature has demonstrated that social-emotional difficulties for children ages 1-2 years old, such as externalizing, internalizing, and dysregulation problems, persist into preschool, especially for at-risk groups (Alink et al., 2006; Briggs-Gowan, Carter, Bosson-Heenan, Guyer, & Horwitz, 2006). By preschool age, an estimated 13-27% of children meet diagnostic criteria for at least one psychiatric disorder (Bufferd, Dougherty, Carlson, & Klein, 2011; Wichstrøm et al., 2012), including externalizing disorders, emotional disorders, and internalizing disorders, in rank order of greatest prevalence. Therefore, understanding factors contributing to mental health problems in infancy would inform interventions promoting child mental health and well-being.

Parents are arguably the most influential factor in infant development. Not surprisingly, parent mental health problems have deleterious effects on children, especially during infancy (Seifer & Dickstein, 2000). Children of parents with mental health problems are more likely to experience language and cognitive development impairments, school problems, and a wide spectrum of mental health problems (Smith, 2004). This is especially true for families experiencing homelessness, which has been associated with increased maternal mental health problems, substance abuse, and exposure to violence (Gewirtz, DeGarmo, Plowman, August, & Realmuto, 2009). Furthermore, infants experiencing poverty and homelessness are among the most vulnerable populations, given the associations between homelessness and poor nutrition, growth, health outcomes, developmental delays, and later school problems (Lieberman & Osofsky, 2009; Madigan, Moran, Schuengel, Pederson, & Otten, 2007; Wood, Valdez, Hayashi, & Shen, 1990). Undoubtedly, homelessness increases the risk for not only parent mental health problems, but also has a greater impact on children’s mental health. Therefore, interventions targeting infant mental health ultimately must incorporate parenting to maximize outcomes.

### Prevention and Intervention

Given the early emergence of mental health problems during infancy, many prevention and treatment interventions have been created. Prevention and treatment efforts aim to reduce the risk, occurrence, and negative impacts of mental health problems (Arango et al., 2018; Organization, 2004). While a majority of the literature examines prevention and treatment interventions separately, in order to most efficiently and effectively maximize optimal mental health, the prevention science and treatment science must co-exist (Weisz, Sandler, Durlak, & Anton, 2005). Numerous systematic reviews and meta-analyses have evaluated the effectiveness of interventions for children. However, significant limitations have not been addressed, including the examination of interventions that focus on parenting, specifically targeting infants and at-risk populations, such as homelessness. Many parenting interventions promote secure infant-caregiver attachment (Bakermans-Kranenburg, Van IJzendoorn, & Juffer, 2005; van IJzendoorn, Juffer, & Duyvesteyn, 1995). However, attachment-based interventions often fail to examine child mental health outcomes, such as internalizing or externalizing behavior problems. Other studies have reviewed parenting interventions targeting child mental health, demonstrating improvements in child behavioral (e.g., conduct problems, noncompliance) and emotional functioning (e.g., anxiety, depression), but are also limited by not specifically examining infancy (Carr, 2014; E. S. Pearl, 2009; Sampers, Anderson, Hartung, & Scambler, 2001; Tully & Hunt, 2016). Similarly, other prevention efforts begin prenatally, such as the Nurse-Family Partnership (Olds, 2006). While this intervention has demonstrated improvements in parent outcomes (e.g., sensitivity, completion of education, finding jobs), there has been less examination of child outcomes (Olds, 2006). Other home-visitation programs examine child outcomes, but are limited by primarily targeting health behaviors (e.g., premature birth, obesity) rather than mental health (Benzies, Magill-Evans, Hayden, & Ballantyne, 2013; Toomey et al., 2019), or do not focus on infancy (Peacock, Konrad, Watson, Nickel, & Muhajarine, 2013; Sweet & Appelbaum, 2004). Lastly, a review by Bayer and colleagues (2009), only examined prevention programs, which neglects the treatment literature. Identifying prevention and treatment parenting interventions for infants is especially important for at-risk families experiencing homelessness, which is associated with higher rates of externalizing problems, internalizing problems, trauma, and maltreatment (Park et al., 2011).

Existing prevention and treatments for child mental health, draw from multiple theoretical backgrounds (e.g., attachment, social learning), utilize a variety of platforms (e.g., in person, internet delivery), and are deliverable in many modalities (e.g., individual, group) of varying length. However, despite the overwhelming literature base for prevention and treatment interventions, there is very little convergence across mental health outcomes, especially for infants. Furthermore, less is known about the extent to which programs target those in greatest need. Despite being long overdue, few studies examine the effectiveness of interventions for children and families experiencing homelessness (Bassuk et al., 2015).

### Current Study

While numerous biological and environmental factors impact infant mental health, parenting style, behaviors, and the quality of parent-child relationship are undoubtably among the most influential (George & Engel, 1980). Although innumerable prevention and treatment programs exist for children and families, there have been no reviews, to our knowledge, examining parent based programs targeting infant mental health outcomes from birth to age 2 (Uylings, 2006). Furthermore, very little is known about prevention and treatment programs for infants and families experiencing homelessness. Examining intervention programs for this vulnerable population is critically important given not only the rise in the number of homeless children and families but also given their well-established higher rates of mental health problems. Additionally, rates of maltreatment and subsequent trauma are significantly higher among families experiencing homelessness (Herbers, Cutuli, Monn, Narayan, & Masten, 2014). Given the well-documented links between maltreatment and mental health outcomes (e.g., internalizing and externalizing behavior problems; Cicchetti, 2013; Leeb, Lewis, & Zolotor, 2011), it is particularly important to identify programs that measure and/or address trauma or, at the very least, have been implemented in populations experiencing trauma. Identifying effective programs targeting child mental health outcomes during the early years of life would identify gaps in the literature for future research to examine and reach populations in greatest need. Thus, the purpose of the current review was to a) systematically examine prevention and treatment interventions targeting infant mental health outcomes (i.e., internalizing behavior problems, externalizing behavior problems, social-emotional development, trauma) in children from birth to age 2 years, b) highlight strengths and limitations of current interventions, and c) identify gaps in the existing literature to inform future mental health intervention science in areas of greatest need, specifically within the context of homelessness.

## Methods

### Retrieval and Selection of Studies

We used electronic databases (i.e., PsychINFO, Google Scholar) to identified peer-reviewed studies of intervention programs developed over the last 30 years using the following keywords: (*prevention* or *treatment* or *program*) and (*infant* or *toddler*) and (*mental health* or *internalizing problems* or *externalizing problems* or *behavior problems* or *anxiety* or *parent program* or *attachment* or *trauma* or *development* or *parent training*). In addition to searching via keywords, searches were also conducted on well-established early intervention programs found in the What Works Clearinghouse database (e.g., The Incredible Years, Triple P, Parent-Child Interaction Therapy, Child-Parent Psychotherapy). The reference sections of all retrieved articles were searched for any other relevant intervention programs.

### Inclusion and Exclusion Criteria

Interventions were included if they were published in a peer-reviewed journal between 1990 and June 2020 and met the following criteria: 1) parent/caregiver prevention or treatment program targeting mental health or attachment, 2) age range of children served in the program had to involve children from birth to 2 years, and 3) at least one outcome measure of infant mental health (i.e., internalizing, externalizing, trauma). Inclusion criteria was partly informed by recommendations for early intervention from a systems perspective (Guralnick, 2011). Programs were excluded if they primarily targeted infant physical health (e.g., feeding, sleeping), were located within schools, hospitals (e.g., neonatal intensive care units) or primary health care settings (e.g., physician’s offices), or if the intervention was not manualized or manuscripts did not provide a detailed program outline for replication purposes. Additionally, case studies, unpublished studies, or interventions only reported at meetings or conferences were also excluded. From over 15,000 citations initially identified, thirty-three programs were identified that met the criteria for this review. All programs were evaluated for level of empirical support specific to infant mental health outcomes based on the typology for classifying interventions by level of scientific evidence outlined by (Brownson, Fielding, & Maylahn, 2009).

### Level of Scientific Evidence

Brownson et al. (2009) typology for classifying interventions by level of scientific evidence includes four categories ranging from least to most effective: Emerging, Promising, Effective, Evidence-based. Additionally, an “Ineffective” category was included, which represents programs demonstrating no improvements. Category was determined by establishing features (e.g., peer review via systematic or narrative review, ongoing work) with special considerations (e.g., external validity, costs and cost-effectiveness, summative evidence of effectiveness). More specifically, these categories were determined to classify the level of evidence specifically for infant mental health outcomes (e.g., externalizing, internalizing, trauma). Notably, infant-caregiver attachment style was not included in the definition of infant mental health. While attachment is an incredibly important construct related to mental health, especially during infancy, the focus of the current review was to examine programs specifically targeting infant mental health as defined by behavioral and emotional manifestations. See Berlin, Zeanah, and Lieberman (2008) and Bakermans-Kranenburg, Van Ijzendoorn, and Juffer (2003) for a review of programs targeting infant attachment. Additionally, when considering the demographic breakdown of the United States homeless population, Hispanic/Latinx and Black or African American individuals are disproportionately represented (Fusaro, Levy, & Shaefer, 2018). As minority groups are largely underrepresented in research, studies conducted within underrepresented, diverse samples was a requirement for effective and evidence based. Programs were classified as ineffective if they met all inclusion criteria, but all peer-reviewed, published studies revealed no improvements in mental health outcomes for infants at post intervention. Emerging programs had to have at least 1 peer-reviewed manuscript evaluating infant mental health at pre and post intervention; it did not have to include an RCT. Programs were classified as promising if they had at least 1 RCT, which included pilot studies, theoretical grounding, and demonstrated pre to post intervention improvements in infant mental health, even if only within one single measure or scale. The effective category was defined by having at least 2 RCTs within underrepresented, diverse samples, demonstrating positive improvements from pre to post intervention for 50% of the infant mental health outcomes across studies. Lastly programs were classified as evidence-based if they met all criteria for an effective intervention *and* included a meta-analysis or review paper, which showed positive findings for infant mental health from pre to post intervention. Given the focus on homelessness for this review, cost-effectiveness was considered, but not required for any category. For example, while groups have been shown to be more cost-effective, groups may not always be a feasible option within homeless shelters.

## Results

### Program Characteristics

The majority of the programs reviewed were prevention interventions (79%; e.g., Attachment and Biobehavioral Catch-Up (ABC), Circle of Security, Family Check-Up). Additionally, 12% were treatment interventions (e.g., Parent-Child Interaction Therapy (PCIT), Infant Behavior Program). Lastly, some programs (9%) were utilized as both prevention and treatment interventions (e.g., Early Pathways, Incredible Years). See Table 1 for information on all reviewed programs, including type of intervention, age range, target population, and other descriptive features; programs are listed alphabetically by program name, with the exception of program adaptations, which are listed alphabetically under the original program name. Furthermore, Table 2 summarizes demographic information about study samples, number of randomized controlled trials (RCT), and infant mental health outcomes.

**Table 1.**
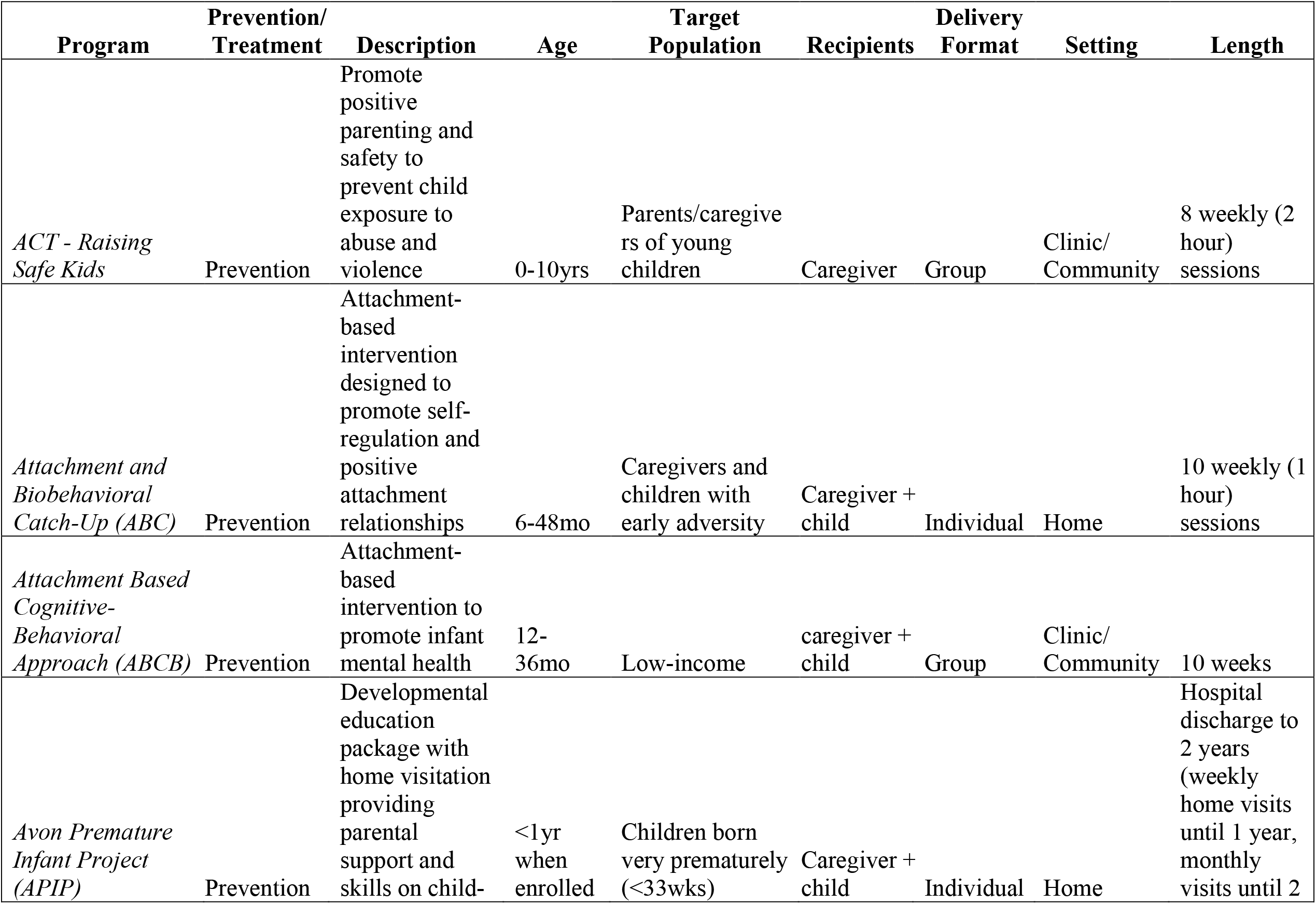
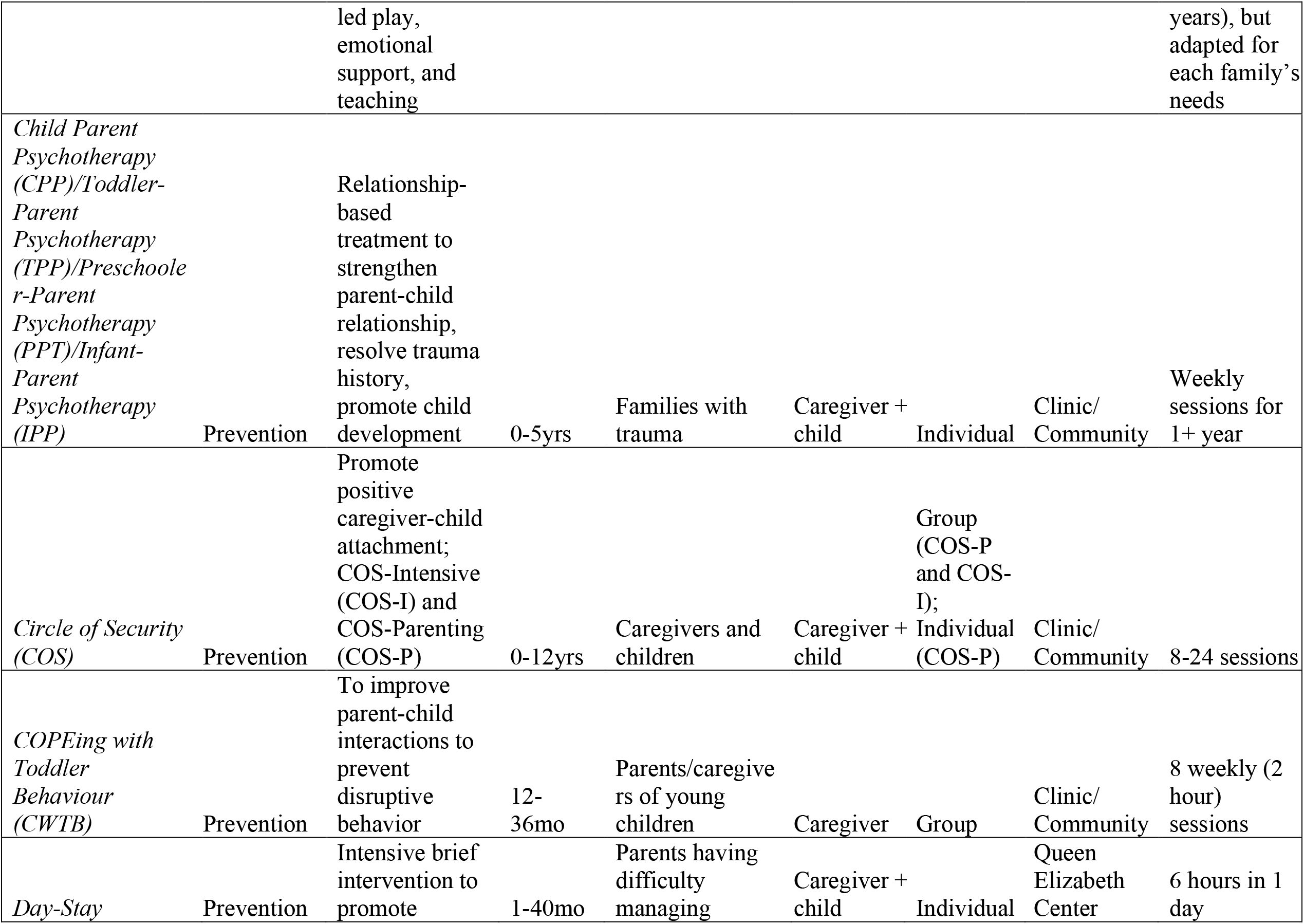
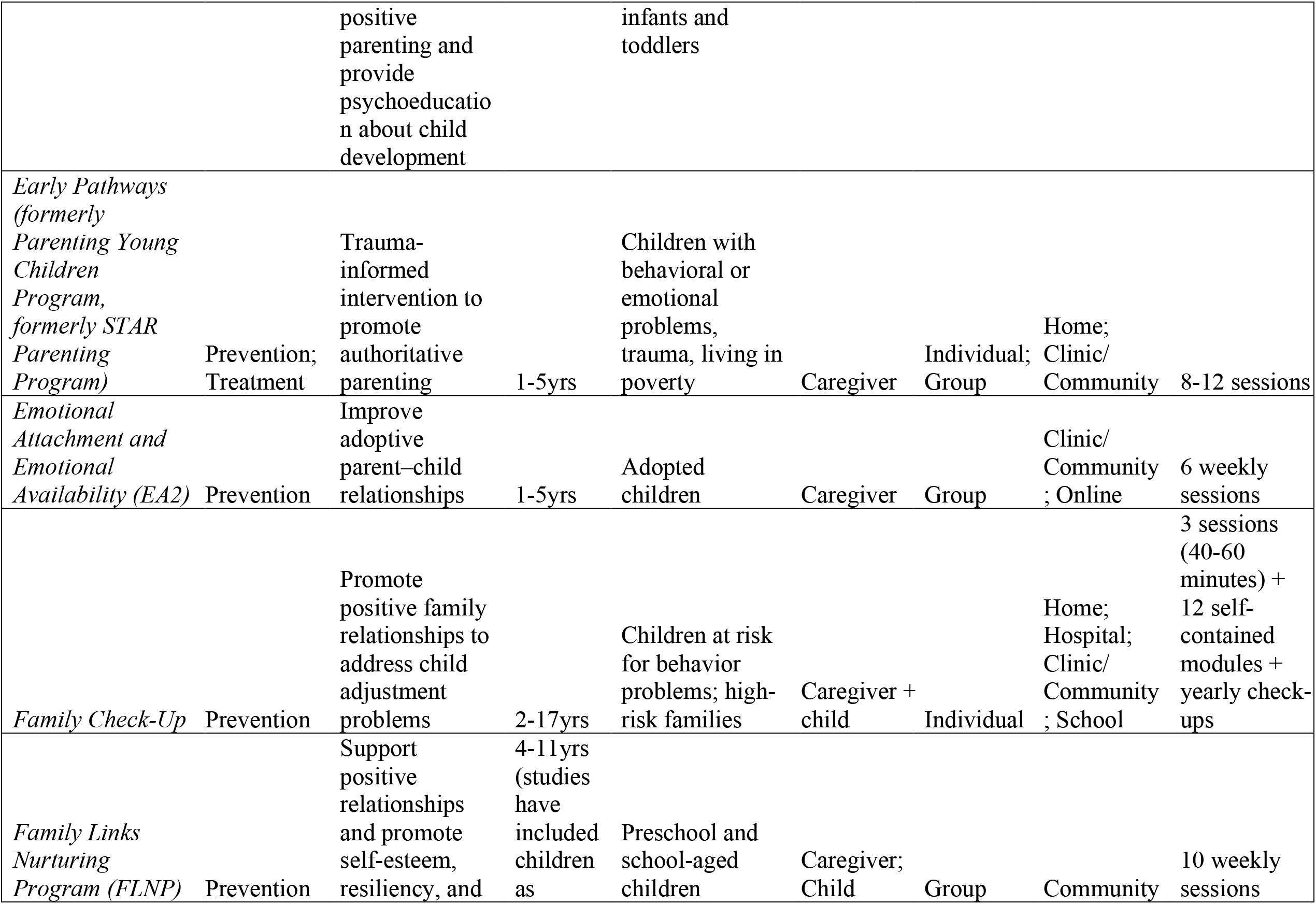
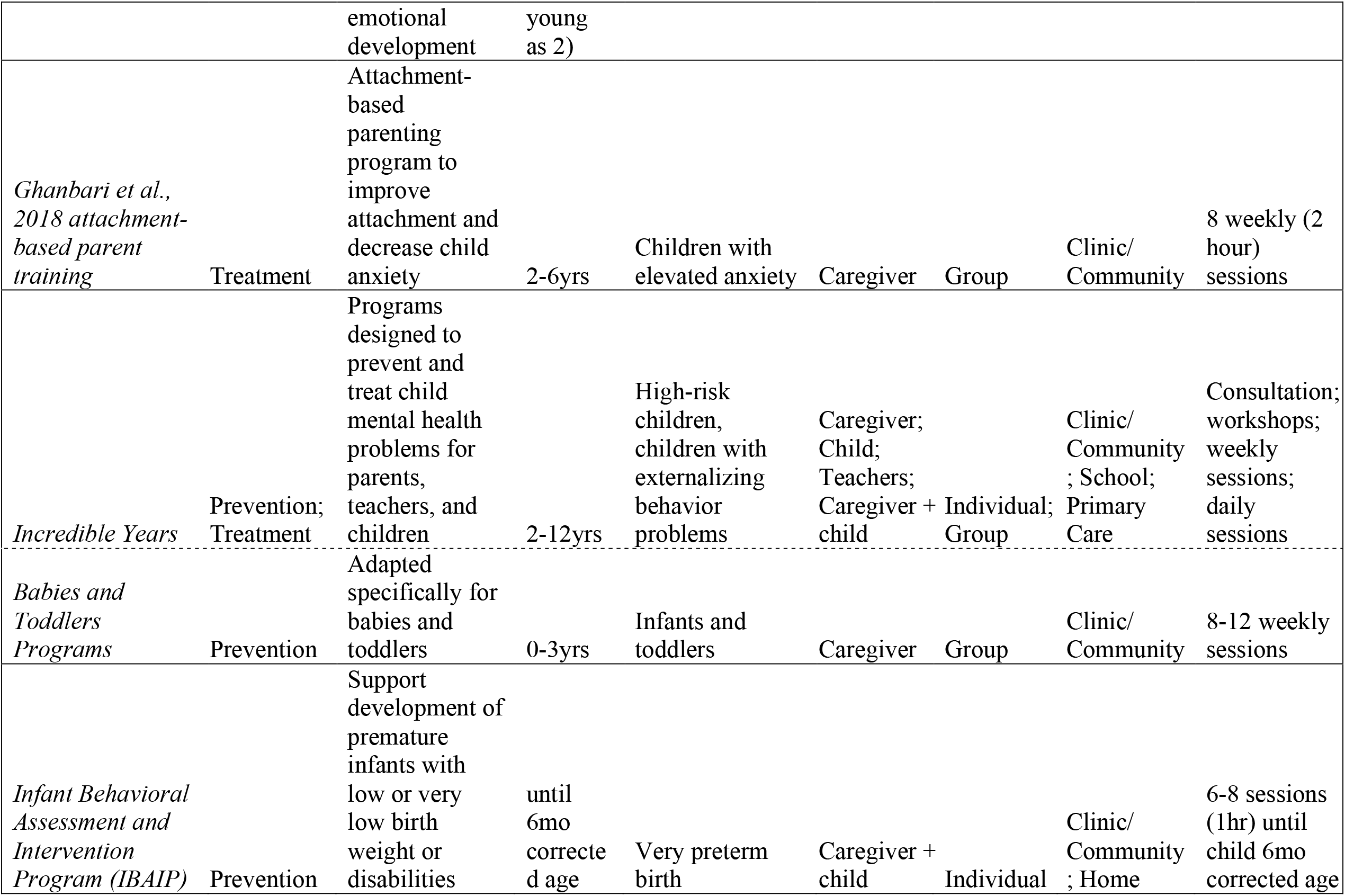
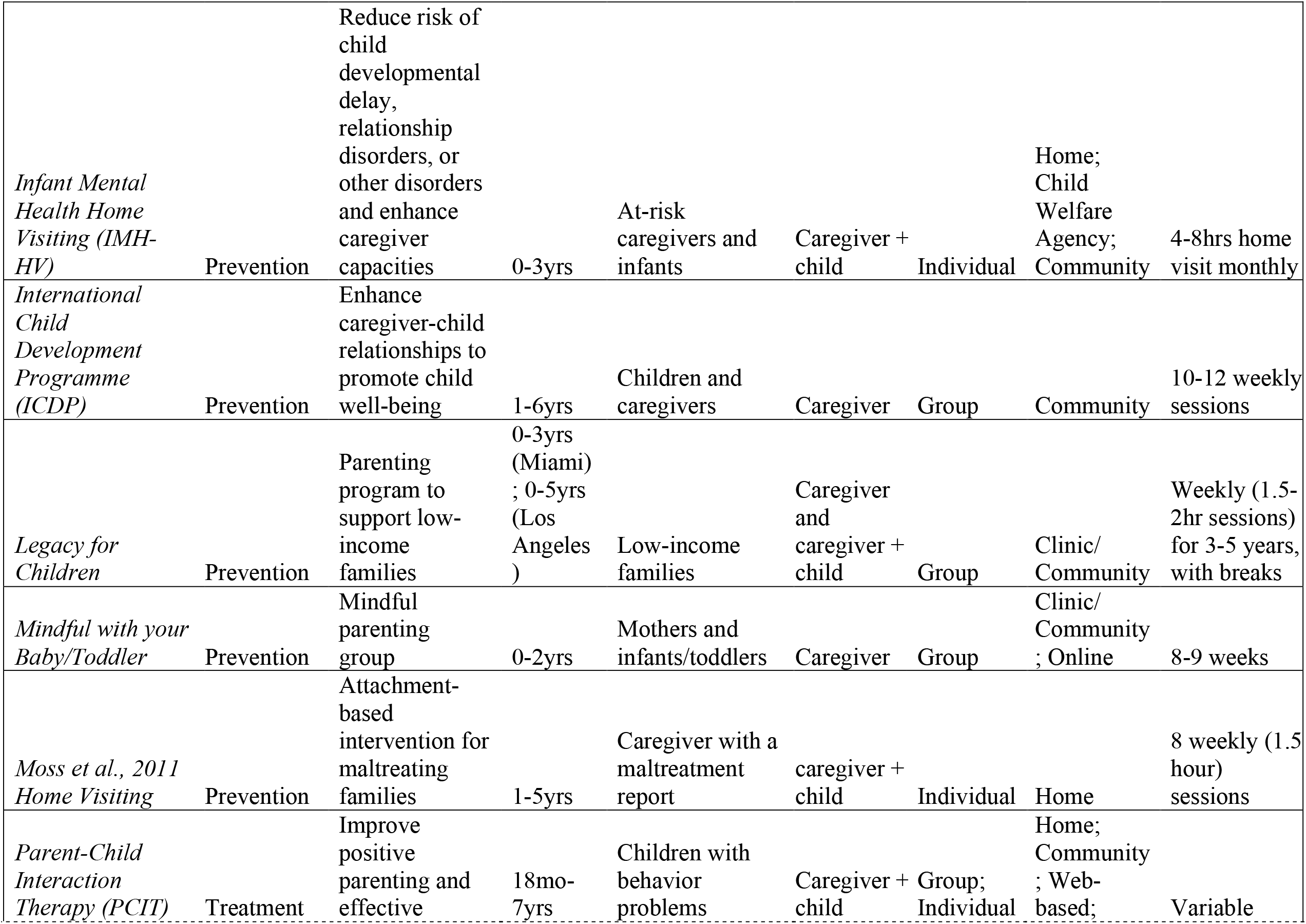
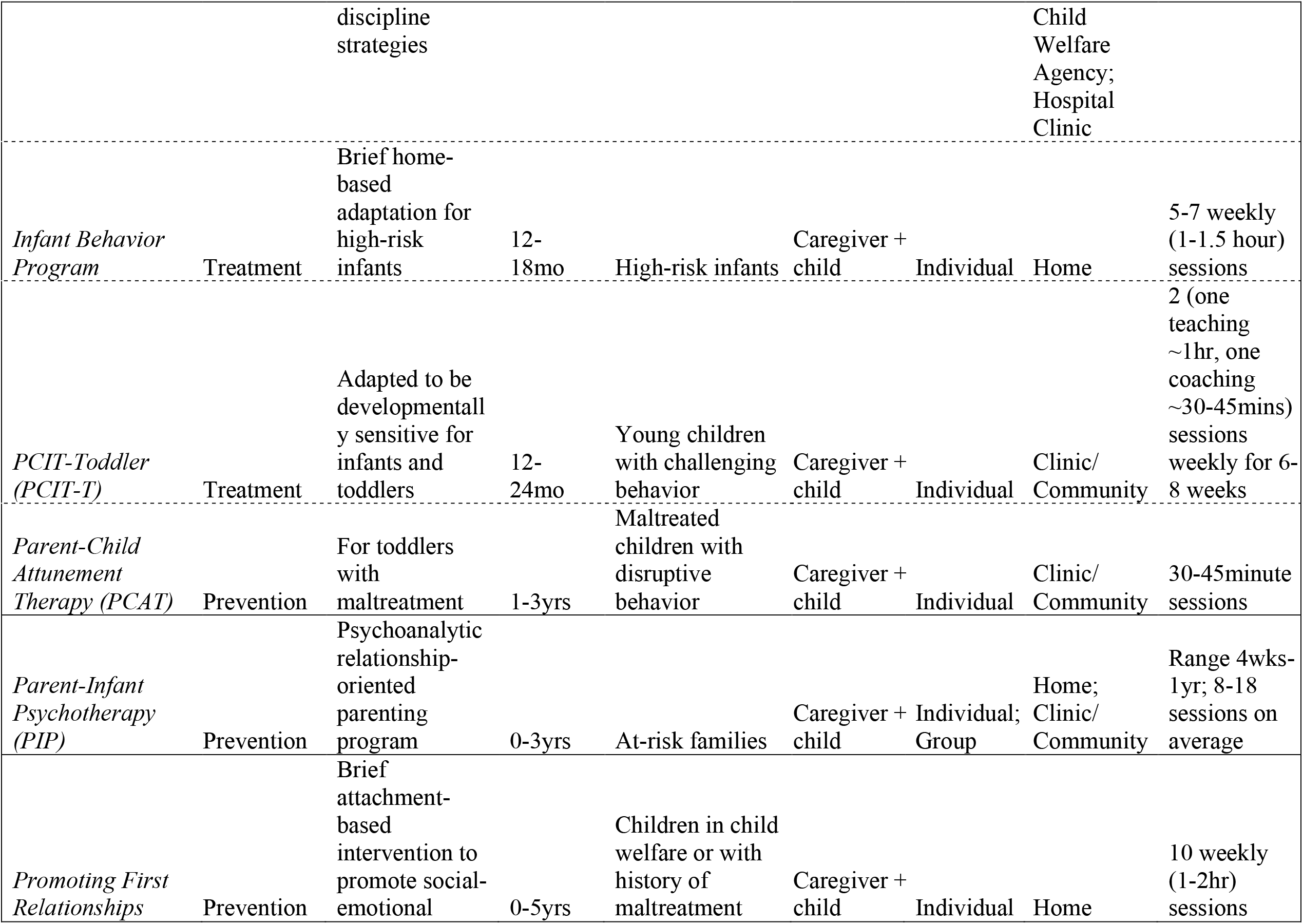
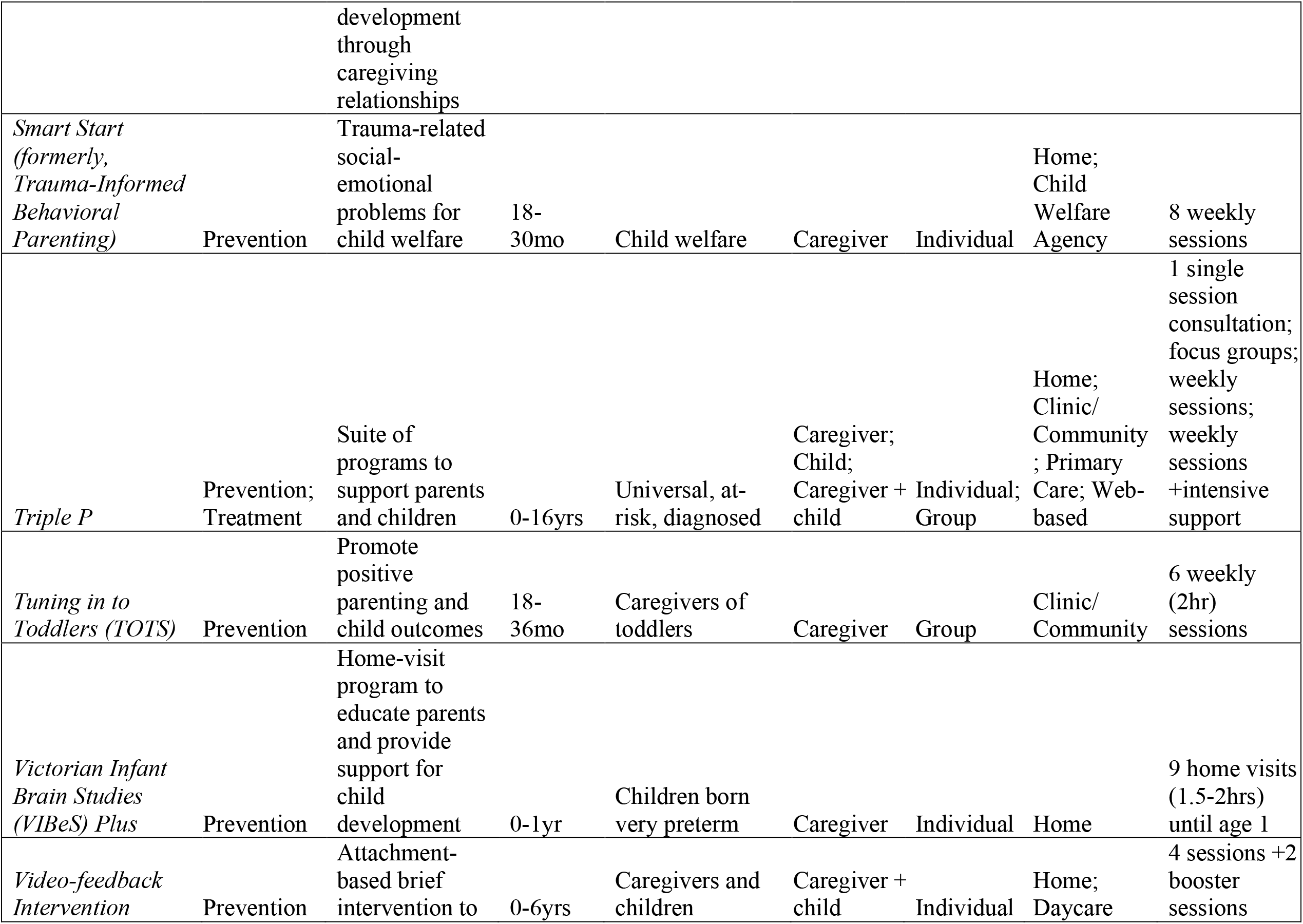
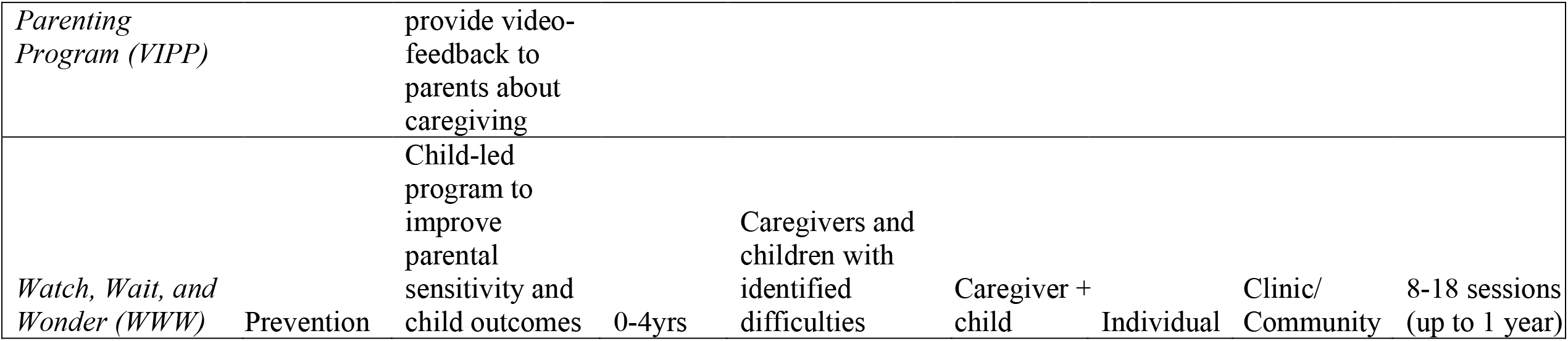
Characteristics of Reviewed Programs

| **Program** | **Prevention/**<br>**Treatment** | **Description** | **Age** | **Target**<br>**Population** | **Recipients** | **Delivery**<br>**Format** | **Setting** | **Length** |
| --- | --- | --- | --- | --- | --- | --- | --- | --- |
| *ACT - Raising Safe Kids* | Prevention | Promote positive parenting and safety to prevent child exposure to abuse and violence | 0-10yrs | Parents/caregivers of young children | Caregiver | Group | Clinic/<br>Community | 8 weekly (2 hour) sessions |
| *Attachment and Biobehavioral Catch-Up (ABC)* | Prevention | Attachment-based intervention designed to promote self-regulation and positive attachment relationships | 6-48mo | Caregivers and children with early adversity | Caregiver + child | Individual | Home | 10 weekly (1 hour) sessions |
| *Attachment Based Cognitive-Behavioral Approach (ABCB)* | Prevention | Attachment-based intervention to promote infant mental health | 12-36mo | Low-income | caregiver + child | Group | Clinic/<br>Community | 10 weeks |
| *Avon Premature Infant Project (APIP)* | Prevention | Developmental education package with home visitation providing parental support and skills on child- | <1yr when enrolled | Children born very prematurely (<33wks) | Caregiver + child | Individual | Home | Hospital discharge to 2 years (weekly home visits until 1 year, monthly visits until 2 |
|  |  | led play, emotional support, and teaching |  |  |  |  |  | years), but adapted for each family's needs |
| *Child Parent Psychotherapy (CPP)/Toddler-Parent Psychotherapy (TPP)/Preschooler-Parent Psychotherapy (PPT)/Infant-Parent Psychotherapy (IPP)* | Prevention | Relationship-based treatment to strengthen parent-child relationship, resolve trauma history, promote child development | 0-5yrs | Families with trauma | Caregiver + child | Individual | Clinic/Community | Weekly sessions for 1+ year |
| *Circle of Security (COS)* | Prevention | Promote positive caregiver-child attachment; COS-Intensive (COS-I) and COS-Parenting (COS-P) | 0-12yrs | Caregivers and children | Caregiver + child | Group (COS-P and COS-I); Individual (COS-P) | Clinic/Community | 8-24 sessions |
| *COPEing with Toddler Behaviour (CWTB)* | Prevention | To improve parent-child interactions to prevent disruptive behavior | 12-36mo | Parents/caregivers of young children | Caregiver | Group | Clinic/Community | 8 weekly (2 hour) sessions |
| *Day-Stay* | Prevention | Intensive brief intervention to promote positive parenting and provide psychoeducation about child development | 1-40mo | Parents having difficulty managing infants and toddlers | Caregiver + child | Individual | Queen Elizabeth Center | 6 hours in 1 day |
| *Early Pathways (formerly Parenting Young Children Program, formerly STAR Parenting Program)* | Prevention; Treatment | Trauma-informed intervention to promote authoritative parenting | 1-5yrs | Children with behavioral or emotional problems, trauma, living in poverty | Caregiver | Individual; Group | Home; Clinic/Community | 8-12 sessions |
| *Emotional Attachment and Emotional Availability (EA2)* | Prevention | Improve adoptive parent-child relationships | 1-5yrs | Adopted children | Caregiver | Group | Clinic/Community; Online | 6 weekly sessions |
| *Family Check-Up* | Prevention | Promote positive family relationships to address child adjustment problems | 2-17yrs | Children at risk for behavior problems; high-risk families | Caregiver + child | Individual | Home; Hospital; Clinic/Community; School | 3 sessions (40-60 minutes) + 12 self-contained modules + yearly check-ups |
| *Family Links Nurturing Program (FLNP)* | Prevention | Support positive relationships and promote self-esteem, resiliency, and emotional development | 4-11yrs (studies have included children as young as 2) | Preschool and school-aged children | Caregiver; Child | Group | Community | 10 weekly sessions |
| *Ghanbari et al., 2018 attachment-based parent training* | Treatment | Attachment-based parenting program to improve attachment and decrease child anxiety | 2-6yrs | Children with elevated anxiety | Caregiver | Group | Clinic/Community | 8 weekly (2 hour) sessions |
| *Incredible Years* | Prevention; Treatment | Programs designed to prevent and treat child mental health problems for parents, teachers, and children | 2-12yrs | High-risk children, children with externalizing behavior problems | Caregiver; Child; Teachers; Caregiver + child | Individual; Group | Clinic/Community; School; Primary Care | Consultation; workshops; weekly sessions; daily sessions |
| *Babies and Toddlers Programs* | Prevention | Adapted specifically for babies and toddlers | 0-3yrs | Infants and toddlers | Caregiver | Group | Clinic/Community | 8-12 weekly sessions |
| *Infant Behavioral Assessment and Intervention Program (IBAIP)* | Prevention | Support development of premature infants with low or very low birth weight or disabilities | until 6mo corrected age | Very preterm birth | Caregiver + child | Individual | Clinic/Community; Home | 6-8 sessions (1hr) until child 6mo corrected age |
| *Infant Mental Health Home Visiting (IMH-HV)* | Prevention | Reduce risk of child developmental delay, relationship disorders, or other disorders and enhance caregiver capacities | 0-3yrs | At-risk caregivers and infants | Caregiver + child | Individual | Home; Child Welfare Agency; Community | 4-8hrs home visit monthly |
| *International Child Development Programme (ICDP)* | Prevention | Enhance caregiver-child relationships to promote child well-being | 1-6yrs | Children and caregivers | Caregiver | Group | Community | 10-12 weekly sessions |
| *Legacy for Children* | Prevention | Parenting program to support low-income families | 0-3yrs (Miami)<br>; 0-5yrs (Los Angeles) | Low-income families | Caregiver and caregiver + child | Group | Clinic/Community | Weekly (1.5-2hr sessions) for 3-5 years, with breaks |
| *Mindful with your Baby/Toddler* | Prevention | Mindful parenting group | 0-2yrs | Mothers and infants/toddlers | Caregiver | Group | Clinic/Community<br>; Online | 8-9 weeks |
| *Moss et al., 2011 Home Visiting* | Prevention | Attachment-based intervention for maltreating families | 1-5yrs | Caregiver with a maltreatment report | caregiver + child | Individual | Home | 8 weekly (1.5 hour) sessions |
| *Parent-Child Interaction Therapy (PCIT)* | Treatment | Improve positive parenting and effective | 18mo-7yrs | Children with behavior problems | Caregiver + child | Group; Individual | Home; Community<br>; Web-based; | Variable |

INFANT MENTAL HEALTH PROGRAMS
|  |  | discipline strategies |  |  |  |  |  | Child Welfare Agency; Hospital Clinic |
| --- | --- | --- | --- | --- | --- | --- | --- | --- |
| *Infant Behavior Program* | Treatment | Brief home-based adaptation for high-risk infants | 12-18mo | High-risk infants | Caregiver + child | Individual | Home | 5-7 weekly (1-1.5 hour) sessions |
| *PCIT-Toddler (PCIT-T)* | Treatment | Adapted to be developmentally sensitive for infants and toddlers | 12-24mo | Young children with challenging behavior | Caregiver + child | Individual | Clinic/Community | 2 (one teaching ~1hr, one coaching ~30-45mins) sessions weekly for 6-8 weeks |
| *Parent-Child Attunement Therapy (PCAT)* | Prevention | For toddlers with maltreatment | 1-3yrs | Maltreated children with disruptive behavior | Caregiver + child | Individual | Clinic/Community | 30-45minute sessions |
| *Parent-Infant Psychotherapy (PIP)* | Prevention | Psychoanalytic relationship-oriented parenting program | 0-3yrs | At-risk families | Caregiver + child | Individual; Group | Home; Clinic/Community | Range 4wks-1yr; 8-18 sessions on average |
| *Promoting First Relationships* | Prevention | Brief attachment-based intervention to promote social-emotional development through caregiving relationships | 0-5yrs | Children in child welfare or with history of maltreatment | Caregiver + child | Individual | Home | 10 weekly (1-2hr) sessions |
| *Smart Start (formerly, Trauma-Informed Behavioral Parenting)* | Prevention | Trauma-related social-emotional problems for child welfare | 18-30mo | Child welfare | Caregiver | Individual | Home; Child Welfare Agency | 8 weekly sessions |
| *Triple P* | Prevention; Treatment | Suite of programs to support parents and children | 0-16yrs | Universal, at-risk, diagnosed | Caregiver; Child; Caregiver + child | Individual; Group | Home; Clinic/Community; Primary Care; Web-based | 1 single session consultation; focus groups; weekly sessions; weekly sessions +intensive support |
| *Tuning in to Toddlers (TOTS)* | Prevention | Promote positive parenting and child outcomes | 18-36mo | Caregivers of toddlers | Caregiver | Group | Clinic/Community | 6 weekly (2hr) sessions |
| *Victorian Infant Brain Studies (ViBeS) Plus* | Prevention | Home-visit program to educate parents and provide support for child development | 0-1yr | Children born very preterm | Caregiver | Individual | Home | 9 home visits (1.5-2hrs) until age 1 |
| *Video-feedback Intervention* | Prevention | Attachment-based brief intervention to | 0-6yrs | Caregivers and children | Caregiver + child | Individual | Home; Daycare | 4 sessions +2 booster sessions |
| *Parenting Program (VIPP)* |  | provide video-feedback to parents about caregiving |  |  |  |  |  |  |
| *Watch, Wait, and Wonder (WWW)* | Prevention | Child-led program to improve parental sensitivity and child outcomes | 0-4yrs | Caregivers and children with identified difficulties | Caregiver + child | Individual | Clinic/Community | 8-18 sessions (up to 1 year) |

**Table 2.**
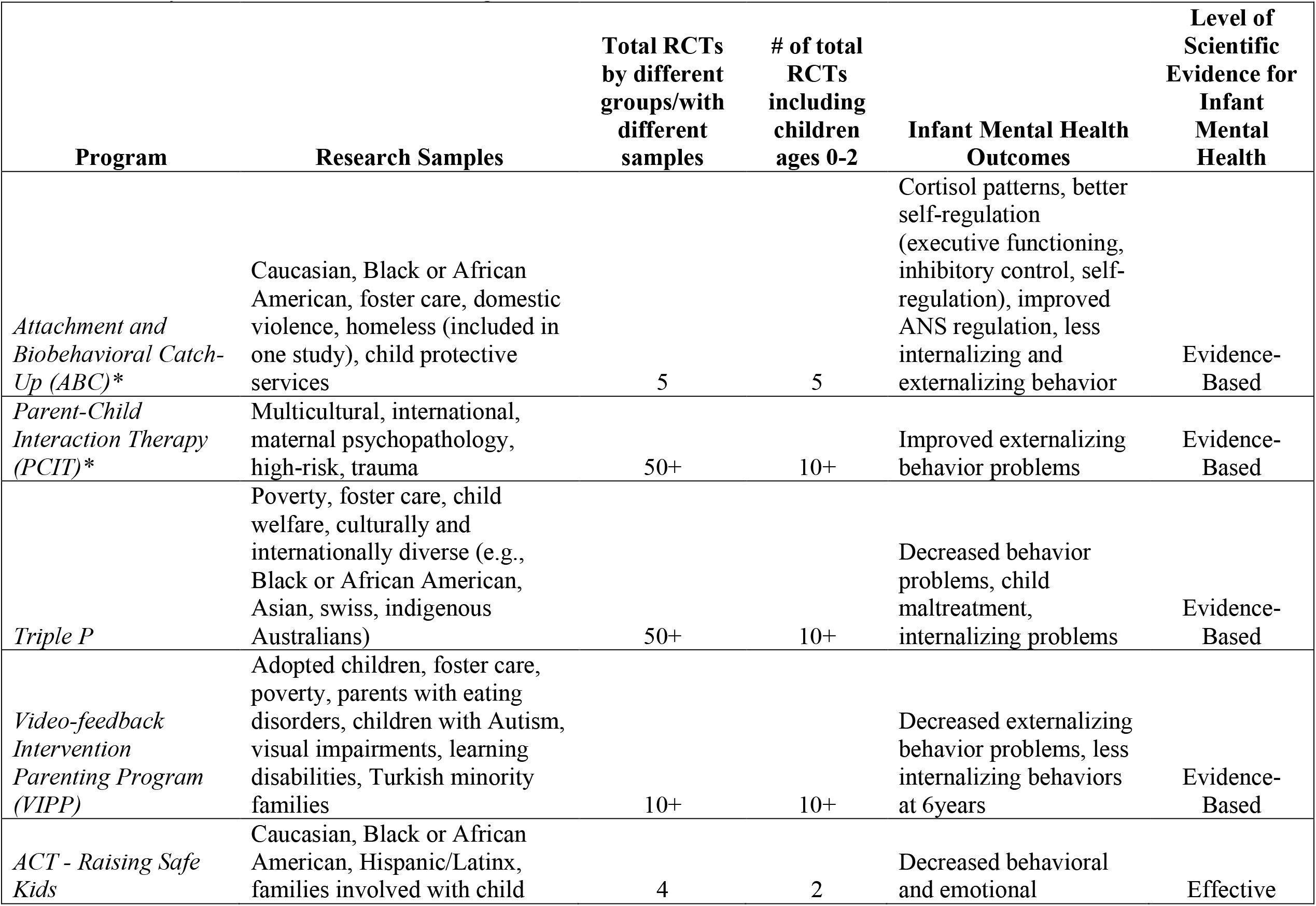
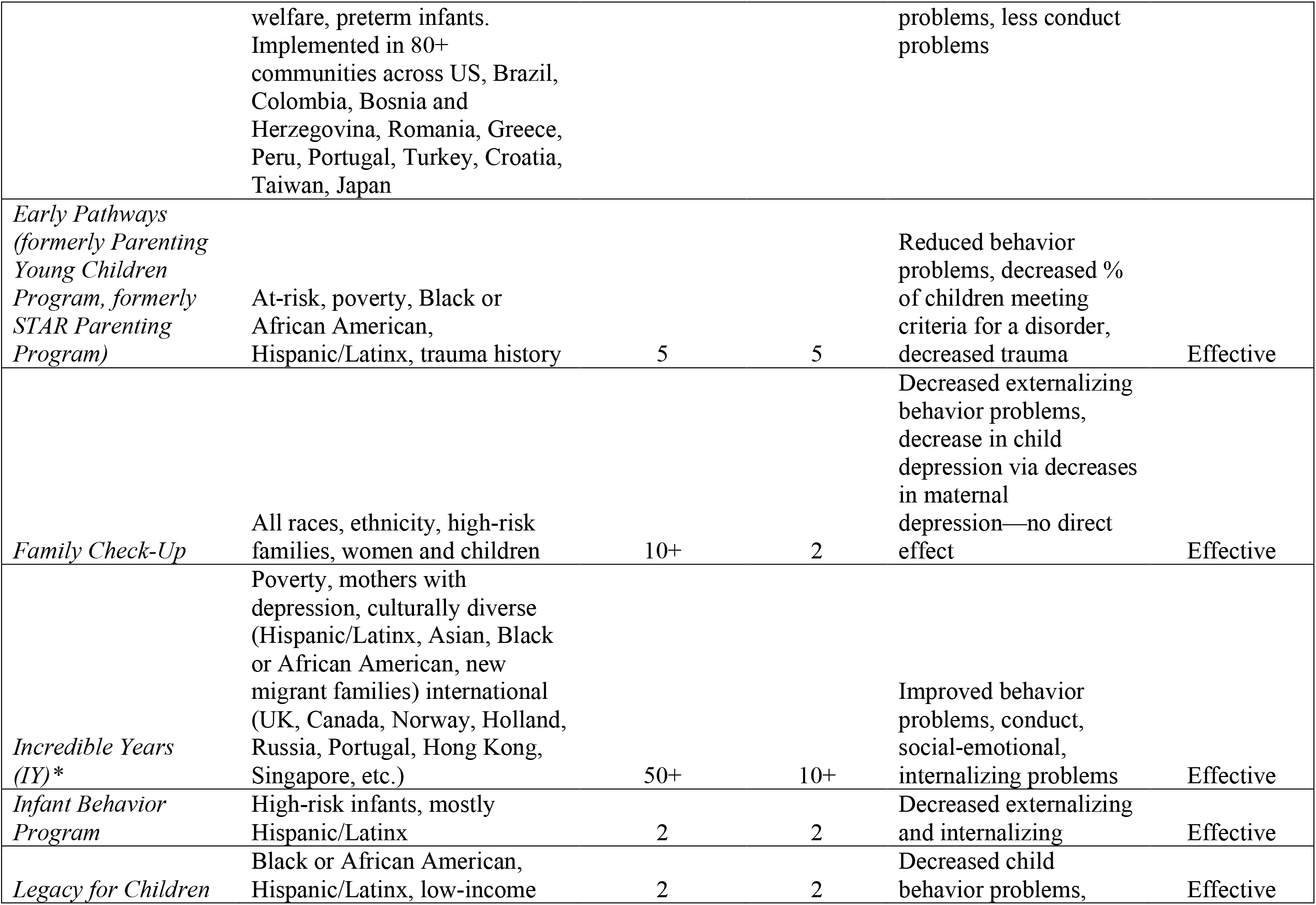
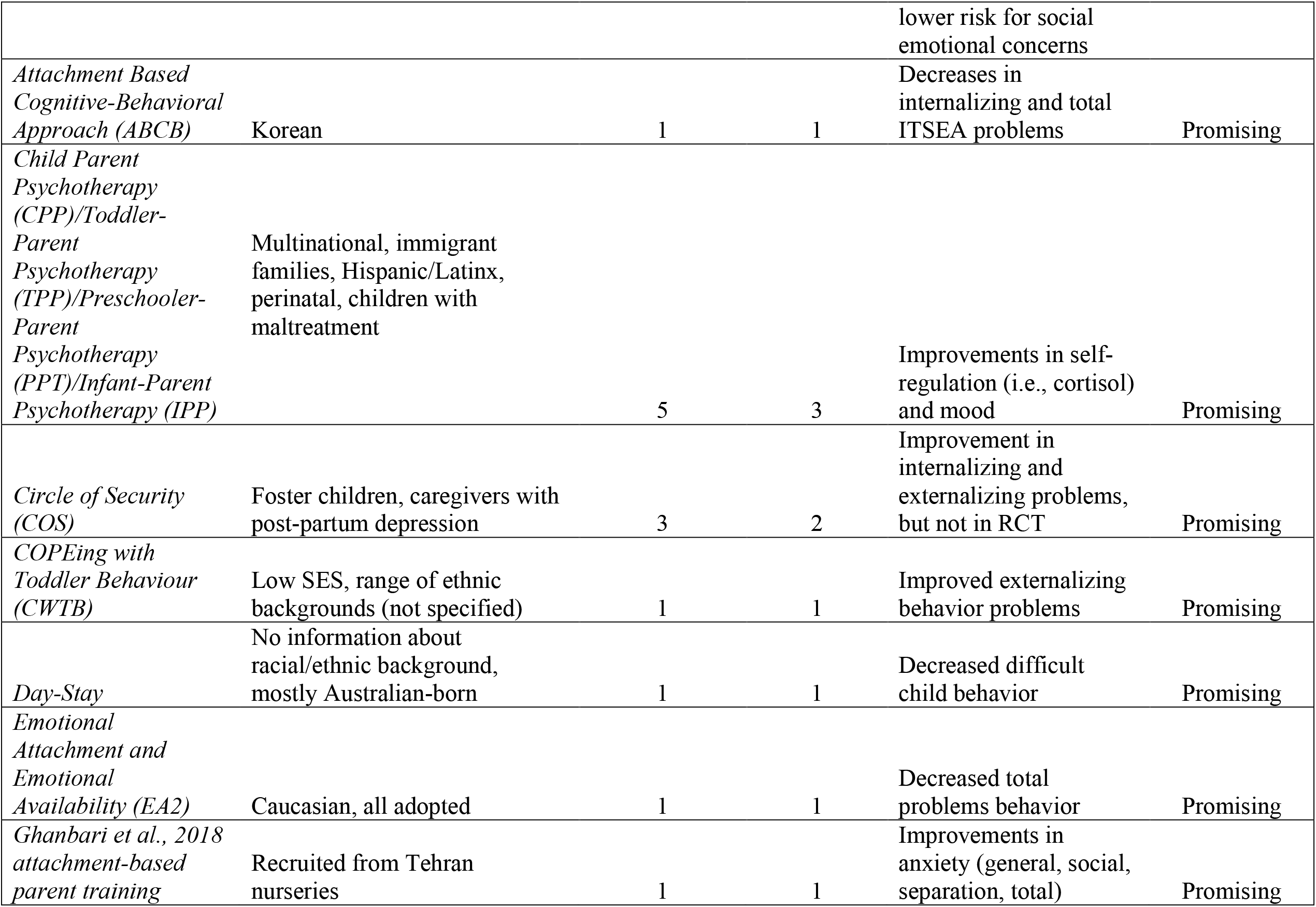
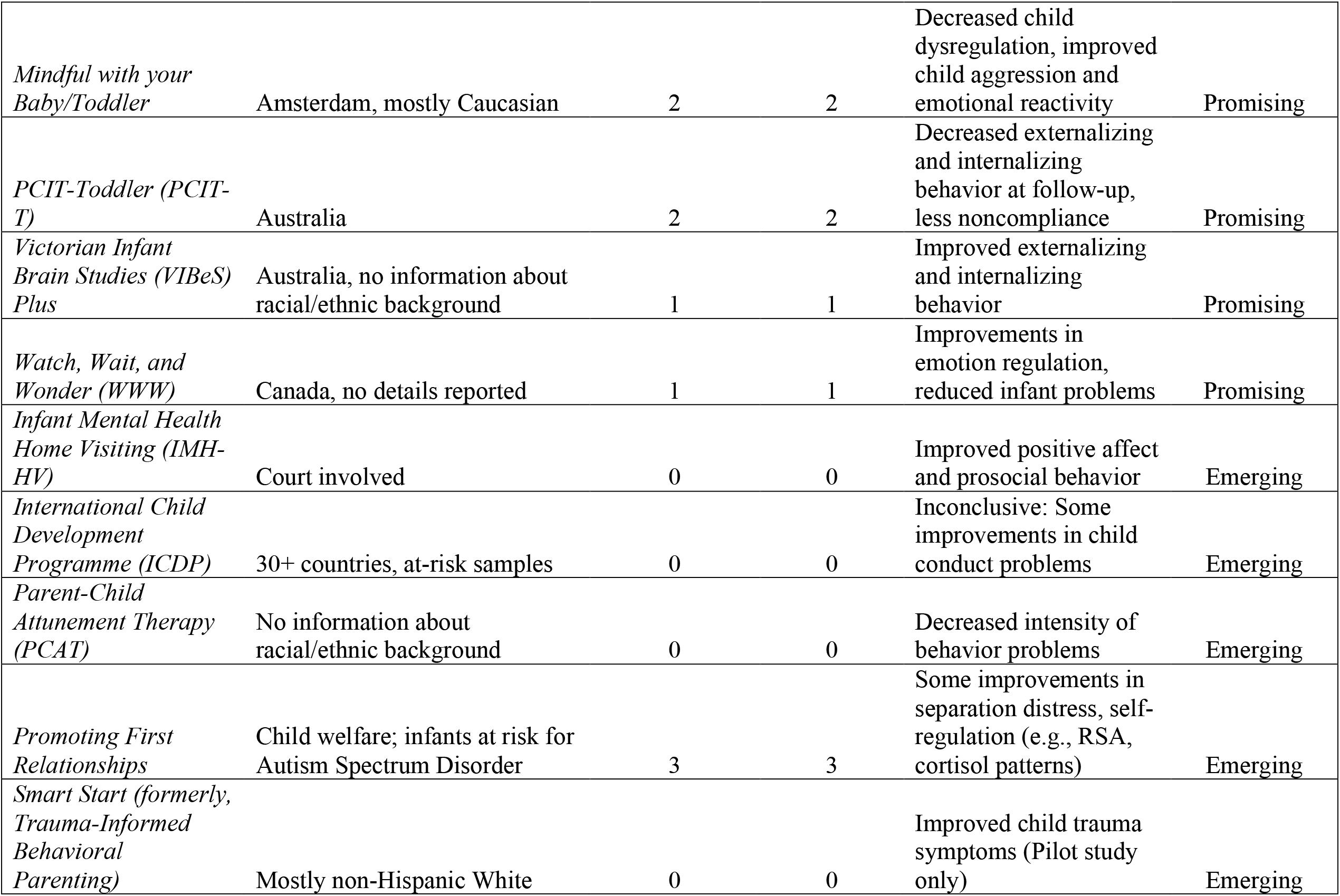
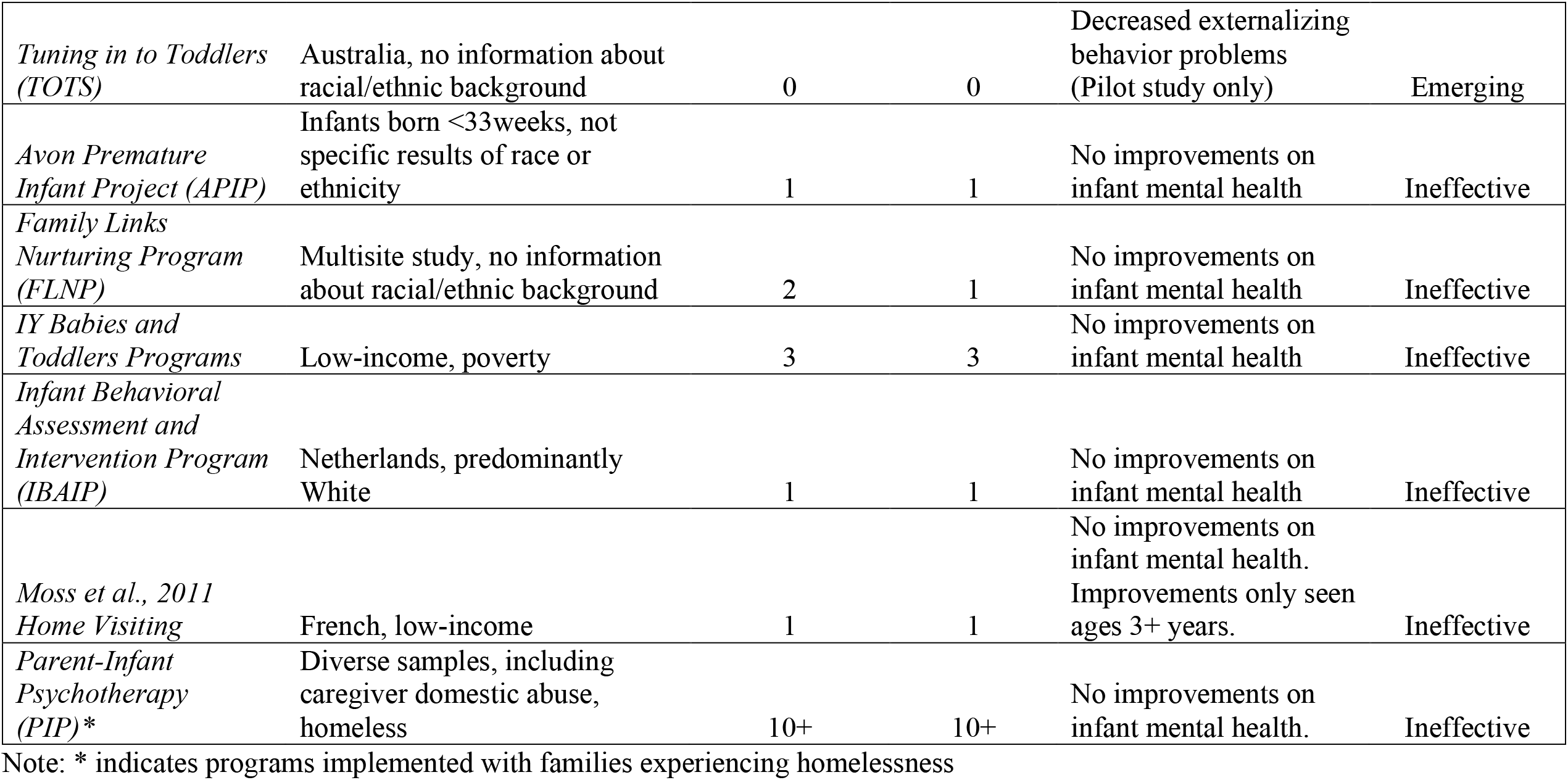
Summary of Evidence for Reviewed Programs

| Program | Research Samples | Total RCTs by different groups/with different samples | # of total RCTs including children ages 0-2 | Infant Mental Health Outcomes | Level of Scientific Evidence for Infant Mental Health |
| --- | --- | --- | --- | --- | --- |
| *Attachment and Biobehavioral Catch-Up (ABC)\** | Caucasian, Black or African American, foster care, domestic violence, homeless (included in one study), child protective services | 5 | 5 | Cortisol patterns, better self-regulation (executive functioning, inhibitory control, self-regulation), improved ANS regulation, less internalizing and externalizing behavior | Evidence-Based |
| *Parent-Child Interaction Therapy (PCIT)\** | Multicultural, international, maternal psychopathology, high-risk, trauma | 50+ | 10+ | Improved externalizing behavior problems | Evidence-Based |
| *Triple P* | Poverty, foster care, child welfare, culturally and internationally diverse (e.g., Black or African American, Asian, swiss, indigenous Australians) | 50+ | 10+ | Decreased behavior problems, child maltreatment, internalizing problems | Evidence-Based |
| *Video-feedback Intervention Parenting Program (VIPP)* | Adopted children, foster care, poverty, parents with eating disorders, children with Autism, visual impairments, learning disabilities, Turkish minority families | 10+ | 10+ | Decreased externalizing behavior problems, less internalizing behaviors at 6years | Evidence-Based |
| *ACT - Raising Safe Kids* | Caucasian, Black or African American, Hispanic/Latinx, families involved with child | 4 | 2 | Decreased behavioral and emotional | Effective |
|  | welfare, preterm infants.<br>Implemented in 80+ communities across US, Brazil, Colombia, Bosnia and Herzegovina, Romania, Greece, Peru, Portugal, Turkey, Croatia, Taiwan, Japan |  |  | problems, less conduct problems |  |
| *Early Pathways (formerly Parenting Young Children Program, formerly STAR Parenting Program)* | At-risk, poverty, Black or African American, Hispanic/Latinx, trauma history | 5 | 5 | Reduced behavior problems, decreased % of children meeting criteria for a disorder, decreased trauma | Effective |
| *Family Check-Up* | All races, ethnicity, high-risk families, women and children | 10+ | 2 | Decreased externalizing behavior problems, decrease in child depression via decreases in maternal depression—no direct effect | Effective |
| *Incredible Years (IY)\** | Poverty, mothers with depression, culturally diverse (Hispanic/Latinx, Asian, Black or African American, new migrant families) international (UK, Canada, Norway, Holland, Russia, Portugal, Hong Kong, Singapore, etc.) | 50+ | 10+ | Improved behavior problems, conduct, social-emotional, internalizing problems | Effective |
| *Infant Behavior Program* | High-risk infants, mostly Hispanic/Latinx | 2 | 2 | Decreased externalizing and internalizing | Effective |
| *Legacy for Children* | Black or African American, Hispanic/Latinx, low-income | 2 | 2 | Decreased child behavior problems, lower risk for social emotional concerns | Effective |
| *Attachment Based Cognitive-Behavioral Approach (ABCB)* | Korean | 1 | 1 | Decreases in internalizing and total ITSEA problems | Promising |
| *Child Parent Psychotherapy (CPP)/Toddler-Parent Psychotherapy (TPP)/Preschooler-Parent Psychotherapy (PPT)/Infant-Parent Psychotherapy (IPP)* | Multinational, immigrant families, Hispanic/Latinx, perinatal, children with maltreatment | 5 | 3 | Improvements in self-regulation (i.e., cortisol) and mood | Promising |
| *Circle of Security (COS)* | Foster children, caregivers with post-partum depression | 3 | 2 | Improvement in internalizing and externalizing problems, but not in RCT | Promising |
| *COPEing with Toddler Behaviour (CWTB)* | Low SES, range of ethnic backgrounds (not specified) | 1 | 1 | Improved externalizing behavior problems | Promising |
| *Day-Stay* | No information about racial/ethnic background, mostly Australian-born | 1 | 1 | Decreased difficult child behavior | Promising |
| *Emotional Attachment and Emotional Availability (EA2)* | Caucasian, all adopted | 1 | 1 | Decreased total problems behavior | Promising |
| *Ghanbari et al., 2018 attachment-based parent training* | Recruited from Tehran nurseries | 1 | 1 | Improvements in anxiety (general, social, separation, total) | Promising |
| *Mindful with your Baby/Toddler* | Amsterdam, mostly Caucasian | 2 | 2 | Decreased child dysregulation, improved child aggression and emotional reactivity | Promising |
| *PCIT-Toddler (PCIT-T)* | Australia | 2 | 2 | Decreased externalizing and internalizing behavior at follow-up, less noncompliance | Promising |
| *Victorian Infant Brain Studies (VIBeS) Plus* | Australia, no information about racial/ethnic background | 1 | 1 | Improved externalizing and internalizing behavior | Promising |
| *Watch, Wait, and Wonder (WWW)* | Canada, no details reported | 1 | 1 | Improvements in emotion regulation, reduced infant problems | Promising |
| *Infant Mental Health Home Visiting (IMH-HV)* | Court involved | 0 | 0 | Improved positive affect and prosocial behavior | Emerging |
| *International Child Development Programme (ICDP)* | 30+ countries, at-risk samples | 0 | 0 | Inconclusive: Some improvements in child conduct problems | Emerging |
| *Parent-Child Attunement Therapy (PCAT)* | No information about racial/ethnic background | 0 | 0 | Decreased intensity of behavior problems | Emerging |
| *Promoting First Relationships* | Child welfare; infants at risk for Autism Spectrum Disorder | 3 | 3 | Some improvements in separation distress, self-regulation (e.g., RSA, cortisol patterns) | Emerging |
| *Smart Start (formerly, Trauma-Informed Behavioral Parenting)* | Mostly non-Hispanic White | 0 | 0 | Improved child trauma symptoms (Pilot study only) | Emerging |
| *Tuning in to Toddlers (TOTS)* | Australia, no information about racial/ethnic background | 0 | 0 | Decreased externalizing behavior problems (Pilot study only) | Emerging |
| *Avon Premature Infant Project (APIP)* | Infants born <33weeks, not specific results of race or ethnicity | 1 | 1 | No improvements on infant mental health | Ineffective |
| *Family Links Nurturing Program (FLNP)* | Multisite study, no information about racial/ethnic background | 2 | 1 | No improvements on infant mental health | Ineffective |
| *IY Babies and Toddlers Programs* | Low-income, poverty | 3 | 3 | No improvements on infant mental health | Ineffective |
| *Infant Behavioral Assessment and Intervention Program (IBAIP)* | Netherlands, predominantly White | 1 | 1 | No improvements on infant mental health | Ineffective |
| *Moss et al., 2011 Home Visiting* | French, low-income | 1 | 1 | No improvements on infant mental health. Improvements only seen ages 3+ years. | Ineffective |
| *Parent-Infant Psychotherapy (PIP)\** | Diverse samples, including caregiver domestic abuse, homeless | 10+ | 10+ | No improvements on infant mental health. | Ineffective |

Note: \* indicates programs implemented with families experiencing homelessness

While all programs reviewed included children ranging from birth to 2 years/24 months of age, only 18% of programs targeted infancy only: Avon Premature Infant Project, Infant Behavioral Assessment and Intervention Program, Mindful with your Baby/Toddler, Infant Behavior Program, PCIT-Toddler, Victorian Infant Brains Studies (VIBeS) Plus. Additionally, among the many programs extending beyond the infancy age range (72%), 4 programs (i.e., Family Check-Up, Ghanbari et al. (2018) attachment-based parent training, Incredible Years BASIC, Family Links Nurturing Program) begin at the upper-bound age (i.e., 2 years) of infancy.

### Ineffective Programs

Based on the criteria, six programs (18%) were classified as ineffective for infant mental health outcomes. These programs included Avon Premature Infant Project, Family Links Nurturing Program, Incredible Years Babies and Toddlers Programs, Infant Behavioral Assessment and Intervention Program, Moss et al. (2011) Home Visiting, and Parent-Infant Psychotherapy.

### Emerging Programs

An additional 18% of programs were classified as emerging. Programs in this category included: Infant Mental Health Home Visiting, International Child Development Programme (ICDP), Parent-Child Attunement Therapy, Promoting First Relationships, Smart Start, and Tuning in to Toddlers. Broadly speaking, programs in this category demonstrated improvements in infant mental health primarily within the externalizing domain (e.g., conduct problems).

In terms of sample diversity, ICDP was the only program that has been implemented across multiple countries. Specific to trauma and homelessness, the majority of programs were implemented in at-risk populations (e.g., child welfare, history of child maltreatment), though Smart Start was the only program directly examining trauma outcomes. Smart Start, which was formerly known as Trauma-Informed Behavioral Parenting, (Agazzi et al., 2019), is an intervention designed to meet the social-emotional needs of toddlers ages 18-30 months in child welfare services. Pilot study results demonstrated reductions in trauma symptoms (Agazzi et al., 2019). However, as there was no comparison group or significant reductions in externalizing behavior problems, future research is warranted. Lastly, no programs in this category have been implemented in shelter settings.

### Promising Programs

Most of the programs reviewed were classified as promising interventions (33%). : Attachment Based Cognitive-Behavioral Approach, Child-Parent Psychotherapy (CPP), Circle of Security, COPEing with Toddler Behaviour, Day-Stay, Emotional Attachment and Emotional Availability, Ghanbari et al., (2018) attachment-based parent training, Mindful with your Baby/Toddler, PCIT-Toddler, VIBeS Plus, and Watch, Wait, and Wonder. However, most programs did not demonstrate improvements across multiple domains of infant mental health and were not examined across diverse samples.

Specific to trauma and homelessness, CPP was the only program in this category targeting infants with a history of trauma (Lieberman, Van Horn, & Ippen, 2005). CPP, which encompasses Toddler-Parent Psychotherapy (Cicchetti, Toth, & Rogosch, 1999), Preschooler-Parent Psychotherapy (PPT), and Infant-Parent Psychotherapy (IPP)—hereby referred to collectively as CPP, is a relationship-based intervention for children ages 0-5 years with a history of trauma or other difficulties. CPP targets the child-parent relationship to promote optimal developmental outcomes for the child, and is based in attachment, cognitive-behavioral, social-learning, and psychodynamic theories. Various treatment strategies are employed including the joint construction of a trauma narrative.

Numerous studies have been conducted examining the effectiveness of CPP across diverse samples, including history of child maltreatment, exposure to domestic violence, immigrant families, and low-income families, evaluating the impacts on caregivers and children. Results demonstrated CPP to be effective in reducing child trauma symptoms and clinical severity of depression and behavior problems for children ages 3-5 years (Ippen, Harris, Van Horn, & Lieberman, 2011; Lieberman, Ippen, & Van Horn, 2006; Lieberman et al., 2005; Toth & Gravener, 2012). While one study examining culturally diverse children ages 0-6 years in foster care found CPP was equally effective across racial and ethnic groups in reducing trauma symptomology, (Weiner, Schneider, & Lyons, 2009), most CPP studies examining the effects on infants are less clear. For example, while CPP improved attachment styles in a sample of infants with a history of maltreatment CPP did not outperform psychoeducation on externalizing or internalizing problems (Cicchetti, Rogosch, Toth, & Sturge-Apple, 2011; Stronach, Toth, Rogosch, & Cicchetti, 2013; Toth, Sturge-Apple, Rogosch, & Cicchetti, 2015). Regarding trauma, not only is CPP trauma-informed, it is also one of very few programs directly assessing child trauma outcomes. Furthermore, to date there are no studies examining the effect of CPP for families experiencing homelessness. Therefore, while CPP is evidence based for children ages 3-5, given its limited consistent positive findings specific to infant mental health, it was categorized as promising.

### Effective Programs

A total of six (18%) programs were classified as effective interventions. These programs included ACT-Raising Safe Kids, Early Pathways, Family Check-Up, Incredible Years (IY), Legacy for Children, and the Infant Behavior Program. All programs demonstrated effectiveness across racially and ethnically diverse samples, including samples with majority Hispanic/Latinx or Black or African American participants.

Regarding trauma, all programs in this category have been implemented in high-risk samples including exposure to trauma, child maltreatment, or involved in child welfare and demonstrated improvements in infant mental health. However, Early Pathways and IY were the only program in this category directly demonstrating improvements in trauma. Early Pathways, which was formerly Parenting Young Children Program and STAR Parenting Program (Fox & Gresl, 2014; Fox & Holtz, 2009; Fox & Nicholson, 2003) has evolved over that past 30+ years into a program for children (ages 1-5 years) designed to promote authoritative parenting that includes cognitive components, positive parenting and effective discipline strategies, and advocacy (http://www.earlypathways.com). Early Pathways is a trauma-informed program based in cognitive behavioral therapy, social learning, and child development. Early Pathways has demonstrated moderate to large effect sizes in decreasing behavior problems, emotional problems, trauma symptoms, and overall likelihood to meet diagnostic criteria for a mental health disorder (Fung & Fox, 2014; Harris, Fox, & Love, 2015; Love & Fox, 2019; Nicholson, Anderson, Fox, & Brenner, 2002). However, Early Pathways has yet to be examined in shelter settings.

IY(Webster-Stratton, Reid, & Stoolmiller, 2008) includes a series of compatible programs (i.e., child programs, parent programs, teacher programs, adjunctive home-visiting services) designed to prevent and treat child (ages 2-12) mental health problems (www.incredibleyears.com). IY is theoretically based in social learning, modeling, and attachment. IY Parents and Babies Program and Toddler Basic Program, while described separately, are collectively part of the Parents, Babies, and Toddlers Series (Webster-Stratton, Reid, & Stoolmiller, 2008). IY Parents and Babies Program targets ages 0-1 years, while the Toddler Basic Program targets ages 1-3 years. Both programs consist of weekly group sessions and promote secure attachment and language development. Additionally, the Toddler Basic Program introduces discipline strategies (Webster-Stratton et al., 2008).

Broadly speaking, IY has demonstrated improvements across domains of child functioning among diverse, multicultural, and international samples. Meta-analyses, reviews, and multiple RCTs have demonstrated IY effectiveness in improving externalizing and internalizing behavior problems, conduct problems, social-emotional development, and academic functioning (Leijten et al., 2018; Morpeth et al., 2017; Pearl, 2009; C. Webster-Stratton & Reid, 2018). However, most studies focus on older children (Menting, de Castro, & Matthys, 2013). While there is substantial support for Incredible Years for children as young as 2 years of age, findings from the Incredible Years Babies and Toddlers Programs are limited to parent outcomes (Evans, Davies, Williams, & Hutchings, 2015; Jones, Erjavec, Viktor, & Hutchings, 2016), thus infant mental health data are still emerging. IY broadly is trauma-informed, such that the programs allow for incorporation of trauma into session components (e.g., trauma narrative with puppets), in addition to demonstrating improvements in child outcomes in samples with exposure to trauma (e.g., domestic abuse, incarcerated parents) (C. Webster-Stratton, 2017).

Additionally, IY was the only program in this category to examine infant mental health in a shelter setting. There is a single case study illustrating the successful application of an IY intervention with a 4-year-old girl and her family in the context of a homeless shelter, which describes qualitative improvements in externalizing behavior problems and trauma symptoms (Williams, 2016). However, the effects of IY on trauma or families experiencing homelessness are yet to be directly assessed for IY beyond a case study. Overall, the IY program is an effective program, although their Babies and Toddlers programs have not demonstrated improvements in infant mental health outcomes and is classified as ineffective.

### Evidence-Based Programs

Based on the criteria defined above, only four (12%) of the programs reviewed were classified as evidence-based interventions for improving infant mental health. Programs included ABC, PCIT, Triple P, and VIPP. Each program is detailed below.

### Attachment and Biobehavioral Catch-Up

The ABC (Dozier et al., 2006) intervention consists of the 10 weekly (1 hour) home-based parent/caregiver coaching sessions to promote infant and toddler self-regulation and secure attachment relationships. ABC was developed to meet the needs of infants experiencing early adversity by targeting secure attachment and healthy biological regulation and is theoretically based in attachment theory in addition to stress neurobiology (Dozier, Roben, Caron, Hoye, & Bernard, 2018).

Since inception, ABC has been implemented with diverse samples (e.g., foster care, children involved with Child Protective Services, children adopted internationally) and disseminated to multiple sites across the United States, including community samples. Additionally, providers are currently utilizing telehealth delivery of ABC, though outcome data are not yet available (www.abcintervention.org). Multiple RCTs with follow-up across different samples and research teams have demonstrated the effectiveness of ABC in improving child mental health outcomes. ABC has been reviewed to demonstrate improvements in child attachment style, self-regulation (e.g., cortisol regulation, emotion-regulation, executive functioning), and both externalizing and internalizing behavior problems (Bernard et al., 2012; Dozier et al., 2006; Grube & Liming, 2018; Lind, Bernard, Ross, & Dozier, 2014; Lind, Raby, Caron, Roben, & Dozier, 2017; Tabachnick, Raby, Goldstein, Zajac, & Dozier, 2019). Regarding trauma, to date no studies have examined the impact of ABC on child trauma symptoms or in a homeless shelter. However, ABC is trauma-informed, including sessions dedicated to addressing caregiver trauma history, and has demonstrated improvements in child mental health among populations with elevated trauma (Dozier et al., 2006; Grube & Liming, 2018). Lastly, families experiencing homelessness have participated in ABC, though researchers have yet to examine the effectiveness of ABC for homelessness specifically.

Providers interested in becoming trained in the ABC intervention must attend an in-person 2-day training and receive a year of biweekly supervision, totaling $7,000, not including laptop and camera equipment required for training (www.abcintervention.org). ABC consists of two separate training programs (i.e., ABC-Infant and ABC-Toddler). Cost-effectiveness studies have not been conducted. There are currently 19 states with ABC providers, in addition to 8 international locations (www.abcintervention.org). Overall, the ABC program is an evidence-based parenting program for high-risk families demonstrating a wide range of positive outcomes for infant mental health.

### Parent-Child Interaction Therapy

Parent-Child Interaction Therapy (PCIT) (Eyberg, Boggs, & Algina, 1995; Zisser & Eyberg, 2010) is a behavioral parent training program originally designed to improve caregiver warmth and limit setting, in addition to firm, clear limit-setting, as a treatment for children with clinically elevated behavior problems. PCIT has been delivered individually, in groups, and online across a variety of settings (e.g., community, hospital clinic, home). PCIT is based in developmental and attachment theories. Therapy is broken down into two phases—child directed interaction (CDI), which teaches positive parenting skills to promote desirable child behavior, and parent directed interaction (PDI), which consists of effective discipline strategies and clear limit-setting to decrease severe child behavior. There is no set number of sessions in traditional PCIT, as the number of treatment sessions required to complete PCIT varies as progression through the program is data-driven and is dependent on parents mastering the necessary skills taught in both phases. However, time-limited adaptations have been evaluated and shown to be as effective (Graziano, Ros-Demarize, & Hare, 2020; Thomas & Zimmer-Gembeck, 2012). Many RCTs, meta-analyses, and reviews have demonstrated the effectiveness of PCIT internationally and multiculturally, in a variety of samples, including racial and ethnic minority groups, and high-risk families, indicating significant decreases in child disruptive behaviors and parental stress, while showing significant improvements in positive parenting and the quality of the parent-child relationship (Cooley, Veldorale-Griffin, Petren, & Mullis, 2014; Thomas, Abell, Webb, Avdagic, & Zimmer-Gembeck, 2017; Ward, Theule, & Cheung, 2016), with persistence of treatment gains up to three years later (Boggs et al., 2005; Hood & Eyberg, 2003). PCIT has also been adapted for high-risk infants (i.e., Infant Behavior Program) (Bagner, Rodríguez, Blake, & Rosa-Olivares, 2013), toddlers with maltreatment history (e.g., Parent Child Attunement Therapy) (Dombrowski, Timmer, Blacker, & Urquiza, 2005), and infants and toddlers with challenging behavior (PCIT-Toddler) (Girard, Wallace, Kohlhoff, Morgan, & McNeil, 2018), all of which have been evaluated for children as young as 12 months. Preliminary outcomes have demonstrated improvements in infant externalizing and internalizing behavior problems and compliance (Bagner et al., 2016; Kohlhoff, Morgan, Briggs, Egan, & Niec, 2020), though more research is warranted.

Regarding trauma, PCIT has also been utilized with trauma survivors, child maltreatment, physical abuse, and in a self-contained domestic violence shelter, (Chaffin et al., 2004; Keeshin, Oxman, Schindler, & Campbell, 2015; Lieneman, Brabson, Highlander, Wallace, & McNeil, 2017). One study also found PCIT effective in reducing children’s trauma symptoms, in addition to behavior problems (Pearl et al., 2012). Similarly, the first phase of PCIT (CDI) has been implemented in homeless shelters (Keeshin et al., 2015), though infant outcomes were not examined. Furthermore, PCIT has not been examined in homeless shelters in any RCTs to date.

In terms of training, PCIT International’s website outlines the various ways in which a clinician can become a PCIT Therapist or PCIT Trainer, both of which require certification (www.pcit.org). Requirements include, graduate education, case experience, training attendance, supervision/consultation, application, and assessments. Initial costs can vary depending on the level of certification, around $5,000 per therapist. Cost-effectiveness examinations have been conducted for PCIT, and have demonstrated that clinic compared to home settings and group compared to individual formats are more cost-effective (French, Yates, & Fowles, 2018; Hare & Graziano, 2020). Overall, PCIT is an evidence-based intervention for improving infant mental health.

### Triple P-Positive Parenting Program

Triple P-Positive Parenting Program (Triple P) (Sanders, 1999), is a suite of programs designed to support parents and children ages 0-16 years. Though Triple P was originally developed from social learning, cognitive behavioral, and developmental theories for at-risk children and parents, there are currently five different levels of Triple P designed to work together and have been implemented as both prevention and treatment interventions in over twenty-five countries. The levels of Triple P include Level 1 (e.g., universal media and communications strategy, Level 2 (e.g., brief consultations about minor concerns), Level 3 (e.g., targeted intervention for children with mild to moderate difficulties), Level 4 (e.g., individual or group intervention for severe difficulties) and Level 5 (e.g., intensive support for complex difficulties; www.triplep.net). There is substantial literature on the effectiveness of Triple P with a variety of samples (e.g., poverty, foster care, child welfare) that are racially and ethnically diverse, including multiple meta-analyses. Triple P has been demonstrated to decrease child externalizing and internalizing behavior problems, as well as reduce rates of child maltreatment (Nowak & Heinrichs, 2008; Prinz, Sanders, Shapiro, Whitaker, & Lutzker, 2009; Sanders et al., 2008; Wiggins, Sofronoff, & Sanders, 2009). Fewer studies include infants or examine the effects of Triple P for infants separately (Wilson et al., 2012), though the results are still evident. One meta-analyses included children as young as birth and ran separate analyses examining the impact of age, indicating a greater effectiveness of Triple P for parents of younger children (Nowak & Heinrichs, 2008). Additionally, Triple P has been implemented and deemed feasible for families with trauma, including an institutional residential setting (Glazemakers & Deboutte, 2013). Additionally, Triple P has been assessed for adaptability, but not implemented, in homeless shelters. Mothers recruited from three shelters in Cape Town, South Africa (Wessels & Ward, 2016) deemed the strategies, materials, and delivery methods acceptable, but raised concerns about time constraints, Internet access, and financial costs. Similarly, Haskett, Armstrong, Neal, and Aldianto (2018) examined parents experiencing homelessness opinions on a Triple P seminar. Sheltered parents rated Triple P more favorably than convenience sample but recommended including more information specific to homelessness (e.g., how to create structure in transition). Outcomes for Triple P in shelter settings have not been documented.

Training in Triple P varies based on level, though typically occurs over one to four days, and include skills-based training requiring a minimum score to be eligible for accreditation (www.triplep.net). In addition to the cost of training, practitioners must also purchase the program materials. Despite initial costs, cost-effectiveness analyses indicated positive results for decreased child abuse and neglect and conduct problems (Foster, Prinz, Sanders, & Shapiro, 2008; Sampaio et al., 2018). Overall, Triple P is an evidence-based intervention, though the Babies and Toddlers programs do not have sufficient evidence.

### Video-feedback Intervention Parenting Program

VIPP (F. E. Juffer, Bakermans-Kranenburg, & Van Ijzendoorn, 2008) is a brief home visiting intervention targeting parental sensitivity to improve outcomes for at-risk children ages 0-6 years. VIPP is based in attachment theory and utilizes video-feedback to provide real-life example to parents. VIPP has been adapted and utilized across multiple countries with a variety of samples (e.g., insecure mothers, preterm infants, adopted children, children with Autism, visually impaired children), and demonstrated effectiveness in improving child behavior problems and cortisol levels, and decreasing risk for internalizing disorders at 6-year follow-up (Juffer, Bakermans*-*Kranenburg, & Van Ijzendoorn, 2018; Klein Velderman et al., 2006; Van Zeijl et al., 2006). Regarding trauma, VIPP has been implemented with children in foster care and examined parental trauma as a potential moderator (Schoemaker et al., 2020; van der Asdonk, Cyr, & Alink, 2020), but no studies have directly assessed child trauma outcomes. Similarly, no studies to date have examined the effectiveness of VIPP for families experiencing homelessness.

Training in VIPP involves a multiple day training, followed by session completion with coaches, presentation of a case example, and certification (www.universiteitleiden.nl). Additionally, home-visitors must serve two or more families per year and receive one hour of coaching each year to maintain certification. Registration costs, which include program manual, training, and supervision, total € 1250, or about $1480 (www.universiteitleiden.nl). No cost-effectiveness information is available at this time. Regardless, VIPP is an evidence-based intervention for infant mental health outcomes with demonstrated improvements in externalizing and internalizing behavior problems across diverse samples.

## Discussion

The purpose of this review was to a) systematically examine prevention and treatment interventions targeting infant mental health outcomes, b) highlight strengths and limitations of current interventions, and c) identify gaps in the existing literature to inform future mental health intervention science for those in greatest need (e.g., dissemination in shelter settings). Results demonstrated that only four programs were categorized as evidence-based for infant mental health outcomes. While many programs demonstrated positive outcomes for children beyond the age of 2, the focus of this review was to examine outcomes specific to infants ages 0-2. Regarding services for high-risk populations, many programs examined infant mental health outcomes in samples with a history of trauma, though few programs (12%) directly assessed trauma outcomes for infants. Lastly, only four programs (ABC, IY, PCIT, PIP) have been implemented with families experiencing homelessness.

Infancy is a developmentally sensitive and critical period in which interventions can mitigate negative biological and environmental impacts on development, including mental health (Uylings, 2006; Knudsen, 2004; George & Engel, 1980). However, disentangling typical and atypical behavior, or differentiating between mental health disorders, early in life remains difficult (Bagner et al., 2012; Foreman, 2015; Szaniecki & Barnes, 2016). Therefore, interventions demonstrating improvements across multiple behavioral and social-emotional domains are critically important. Among the programs reviewed in this manuscript, some programs (e.g., ACT-Raising Safe Kids, COPEing with Toddler Behaviour, Ghanbari et al., 2018 attachment-based parent training, Tuning Into Tots) only demonstrated improvements in one domain of infant mental health. Alternatively, programs such as ABC, Early Pathways, IY, PCIT and adaptations, Triple P, and VIPP demonstrated improvements across multiple domains of infant mental health. However, a large portion of the literature base for IY has not demonstrated robust impacts on mental health for infants younger than 2 years of age. Therefore, such programs may not be appropriate for younger infants.

Additionally, while many programs have demonstrated improvements in infant mental health in high-risk samples (e.g., foster care, low income), almost none of the programs reviewed have been implemented with families experiencing homelessness. Even among programs implemented with families in shelter settings, none have evaluated the effects for infant mental health with a RCT. For IY, qualitative improvements have been published, though it was in the form of a single case study and the child was 4 years old (Williams, 2016). PCIT is the only program with documented improvements specifically within homeless shelters (Keeshin et al., 2015), though never in the context of a RCT. Lastly, Triple P has been evaluated for feasibility in shelter settings (Haskett et al., 2018; Wessels & Ward, 2016), but outcomes related to implementation have not been reported. Examining the feasibility and efficacy of parenting programs for families experiencing homelessness in terms of promoting infant mental health remains a critical and understudied area for research.

It is important to note that while this study is among the first to examine parenting interventions targeting infant mental health, some limitations should be addressed. First and foremost, the findings of this review should be interpreted with caution regarding overall public health impact. As outlined by the RE-AIM framework (Glasgow, Vogt, & Boles, 1999), an implementation science model, an intervention’s public health impact is a function of reach, efficacy, adoption, implementation, and maintenance. While we considered as many factors as possible when determining the level of scientific evidence for each program, we were unable to take into consideration all five factors for each intervention. In terms of program reach, we found no specific participation/eligibility statistics for any program included in this review. When examining efficacy, the current review focused more on positive outcomes, and less on negative outcomes of infant mental health. Additionally, many programs did not utilize multi-informant or multimethod assessments to examine infant mental health outcomes, which limits generalizability of findings. For program adoption, most programs categorized as effective or evidence-based did demonstrate feasibility and acceptability. However, specific to homelessness, almost none of the programs reported adoption statistics. This is an especially important area for future research to examine given the unique barriers to service specific to shelter settings, such as affordability of services and lack of transportation (Kushel, 2015). Implementation was heavily considered in this review, such that effective and evidence-based interventions demonstrated improvements in diverse samples, given that Hispanic/Latinx and Black or African American individuals are disproportionately represented in homeless shelters (Fusaro et al., 2018). Additionally, many interventions were evaluated in community or home settings, supporting dissemination; although it is important to note that most studies were conducted in the context of research. One large limitation of parenting interventions broadly is high rates of attrition (Chacko et al., 2016), and the programs reviewed in this manuscript are no exception. Lastly, given the dearth of literature specific to interventions promoting infant mental health at follow-up time-points, we were unable to compare maintenance of gains between programs. While some programs evaluated follow-up effects, many programs did not; therefore, the results presented in this review were specifically for pre to post intervention outcomes. However, with the emergence of novel infant mental health programs, it will be important for future research to collect follow-up data to determine maintenance effects.

The limitations of the current study highlight the need for policy change and future research. The World Association for Infant Mental Health recently published a task force report examining the burden of mental health during infancy (Lyons-Ruth et al., 2017). This report established global priorities to address infant mental health which include 1) global education about signs of disorder in infancy and toddlerhood, 2) enhancing intervention availability for infants and caregivers, and 3) developing infant and toddler mental health data for developing and war-torn countries. The findings of this review align with the goal in enhancing intervention availability, and further support the need for future work on dissemination of interventions for infant mental health, specifically to vulnerable populations such as families experiencing homelessness. Given the many complexities of shelter settings, including lack of transportation, funding agencies and policymakers should consider embedding programs within the shelter setting. The shelter system already embeds physical health, social work, and assists families with food, employment, and housing, as well as caregiver mental health, such as substance use (Substance Abuse and Mental Health Services Administration, 2020; Kushel, 2015). However, parenting programs are currently not embedded within shelter settings, an area for future policy given the added parent and infant mental health stressors that occur in this setting (Gewirtz et al., 2009; Lieberman & Osofsky, 2009). Lastly, when considering programs for implementation in homeless shelters, length of program and delivery method are important factors given high turnover rates in shelter settings (U.S. Department of Housing and Urban Development, 2012). For example, programs like ABC, which is only 10 weeks long, may be more feasible in this setting. Additionally, one current RCT study examined the feasibility of 12 session time-limited CPP versus PCIT within a homeless shelter. This study, which included children as young as 18 months, found that time-limited PCIT resulted in greater reductions in maternal negative verbalizations and parenting stress, and greater increases in maternal positive verbalizations relative to time-limited CPP. At the child-level, both PCIT and CPP resulted in significant decreases in children’s post-traumatic stress symptoms; however, only PCIT resulted in significant improvements in behavior problems. Taken together these results indicate that although time-limited PCIT and CPP offer effective intervention for children and families experiencing homelessness, PCIT provides a more comprehensive and transdiagnostic service (Graziano et al., under review).

It is abundantly clear from the current review that there are very few interventions with scientific evidence promoting infant mental health. Among populations at greatest risk for mental health problems (i.e., homeless), almost no studies have examined the effectiveness of parenting programs, with no RCT studies to date demonstrating improvements in infant mental health for families in homeless shelters. This review has highlighted the need for future research examining programs for infant mental health, specifically within shelters. While current infrastructure provides physical health care, job training, and food to families in shelters, the same importance has not been placed on infant mental health. It is our view that providing *comprehensive* infant mental health programs, not just developmental screeners, is equally as important. Programs targeting parenting have the potential to mitigate detrimental lifelong impacts of the social-emotional and behavioral difficulties associated with the homeless experience.

## Data Availability

this is a review paper and thus no raw data is available to share.

## Declarations

### Funding and Conflicts of Interest

This work was supported by the National Institute of Mental Health, [grant R01MH112588] and the National Institute on Drug Abuse [T32DA043449]. The opinions expressed are those of the authors and do not represent views of NIH. We would like to acknowledge the support of Sundari Foundation, Inc. dba Lotus House Women’s Shelter (Lotus House) who inspired us to conduct this review.

### Author Contributions

All authors contributed to the manuscript. Paulo Graziano was primarily responsible for the idea for the article, while Taylor Landis and Megan Hare performed the literature search and data analysis. Taylor Landis drafted the manuscript, which was critically revised by both Paulo Graziano and Megan Hare. All authors read and approved the final manuscript.

## Notes

### Competing Interest Statement

The authors have declared no competing interest.

### Clinical Trial

n/a

### Author Declarations

IRB is exempt as this is a review paper

## References

Agazzi, H., Adams, C., Ferron, E., Ferron, J., Shaffer-Hudkins, E., & Salloum, A. (2019). Trauma-Informed behavioral parenting for early intervention. Journal of Child and Family Studies, 28(8), 2172–2186.

Alink, L. R., Mesman, J., Van Zeijl, J., Stolk, M. N., Juffer, F., Koot, H. M., … Van IJzendoorn, M. H. (2006). The early childhood aggression curve: Development of physical aggression in 10-to 50-month-old children. Child development, 77(4), 954–966.

Arango, C., Díaz-Caneja, C. M., McGorry, P. D., Rapoport, J., Sommer, I. E., Vorstman, J. A., … Freedman, R. (2018). Preventive strategies for mental health. The Lancet Psychiatry, 5(7), 591–604.

Bagner, D. M., Coxe, S., Hungerford, G. M., Garcia, D., Barroso, N. E., Hernandez, J., & Rosa-Olivares, J. (2016). Behavioral parent training in infancy: A window of opportunity for high-risk families. Journal of abnormal child psychology, 44(5), 901–912.

Bagner, D. M., Rodríguez, G. M., Blake, C. A., Linares, D., & Carter, A. S. (2012). Assessment of behavioral and emotional problems in infancy: A systematic review. Clinical child and family psychology review, 15(2), 113–128.

Bagner, D. M., Rodríguez, G. M., Blake, C. A., & Rosa-Olivares, J. (2013). Home-based preventive parenting intervention for at-risk infants and their families: An open trial. Cognitive and Behavioral Practice, 20(3), 334–348.

Bakermans-Kranenburg, M. J., Van Ijzendoorn, M. H., & Juffer, F. (2003). Less is more: meta-analyses of sensitivity and attachment interventions in early childhood. Psychological bulletin, 129(2), 195.

Bakermans-Kranenburg, M. J., Van IJzendoorn, M. H., & Juffer, F. (2005). Disorganized infant attachment and preventive interventions: A review and meta-analysis. Infant Mental Health Journal: Official Publication of The World Association for Infant Mental Health, 26(3), 191–216.

Bassuk, E. L., Buckner, J. C., Perloff, J. N., & Bassuk, S. S. (1998). Prevalence of mental health and substance use disorders among homeless and low-income housed mothers. American Journal of Psychiatry, 155(11), 1561–1564.

Bassuk, E. L., Richard, M. K., & Tsertsvadze, A. (2015). The prevalence of mental illness in homeless children: A systematic review and meta-analysis. Journal of the American Academy of Child & Adolescent Psychiatry, 54(2), 86-96. e82.

Bayer, J., Hiscock, H., Scalzo, K., Mathers, M., McDonald, M., Morris, A., … Wake, M. (2009). Systematic review of preventive interventions for children’s mental health: what would work in Australian contexts? Australian & New Zealand Journal of Psychiatry, 43(8), 695–710.

Benzies, K. M., Magill-Evans, J. E., Hayden, K. A., & Ballantyne, M. (2013). Key components of early intervention programs for preterm infants and their parents: a systematic review and meta-analysis. BMC pregnancy and childbirth, 13(S1), S10.

Berlin, L. J., Zeanah, C. H., & Lieberman, A. F. (2008). Prevention and intervention programs for supporting early attachment security.

Bernard, K., Dozier, M., Bick, J., Lewis-Morrarty, E., Lindhiem, O., & Carlson, E. (2012). Enhancing attachment organization among maltreated children: Results of a randomized clinical trial. Child development, 83(2), 623–636.

Boggs, S. R., Eyberg, S. M., Edwards, D. L., Rayfield, A., Jacobs, J., Bagner, D., & Hood, K. K. (2005). Outcomes of parent-child interaction therapy: A comparison of treatment completers and study dropouts one to three years later. Child & Family Behavior Therapy, 26(4), 1–22.

Briggs-Gowan, M. J., Carter, A. S., Bosson-Heenan, J., Guyer, A. E., & Horwitz, S. M. (2006). Are infant-toddler social-emotional and behavioral problems transient? Journal of the American Academy of Child & Adolescent Psychiatry, 45(7), 849–858.

Brownson, R. C., Fielding, J. E., & Maylahn, C. M. (2009). Evidence-based public health: a fundamental concept for public health practice. Annual review of public health, 30, 175–201.

Bufferd, S. J., Dougherty, L. R., Carlson, G. A., & Klein, D. N. (2011). Parent-reported mental health in preschoolers: findings using a diagnostic interview. Comprehensive psychiatry, 52(4), 359–369.

Carr, A. (2014). The evidence base for family therapy and systemic interventions for child-focused problems. Journal of Family Therapy, 36(2), 107–157.

Carr, A. (2019). Family therapy and systemic interventions for child-focused problems: The current evidence base. Journal of Family Therapy, 41(2), 153–213.

Carter, A. S., Briggs-Gowan, M. J., & Davis, N. O. (2004). Assessment of young children’s social-emotional development and psychopathology: Recent advances and recommendations for practice. Journal of Child Psychology and Psychiatry, 45(1), 109–134.

Chacko, A., Jensen, S. A., Lowry, L. S., Cornwell, M., Chimklis, A., Chan, E. …, & Pulgarin, B. (2016). Engagement in behavioral parent training: Review of the literature and implications for practice. Clinical Child and Family Psychology Review, 19(3), 204–215.

Chaffin, M., Silovsky, J. F., Funderburk, B., Valle, L. A., Brestan, E. V., Balachova, T., … Bonner, B. L. (2004). Parent-child interaction therapy with physically abusive parents: efficacy for reducing future abuse reports. Journal of consulting and clinical psychology, 72(3), 500.

Cicchetti, D. (2013). Annual research review: Resilient functioning in maltreated children–past, present, and future perspectives. Journal of Child Psychology and Psychiatry, 54(4), 402–422.

Cicchetti, D., Rogosch, F. A., Toth, S. L., & Sturge-Apple, M. L. (2011). Normalizing the development of cortisol regulation in maltreated infants through preventive interventions. Development and psychopathology, 23(3), 789.

Cicchetti, D., Toth, S. L., & Rogosch, F. A. (1999). The efficacy of toddler-parent psychotherapy to increase attachment security in offspring of depressed mothers. Attachment & Human Development, 1(1), 34–66.

Cooley, M. E., Veldorale-Griffin, A., Petren, R. E., & Mullis, A. K. (2014). Parent–Child Interaction Therapy: A meta-analysis of child behavior outcomes and parent stress. Journal of Family Social Work, 17(3), 191–208.

Dombrowski, S. C., Timmer, S. G., Blacker, D. M., & Urquiza, A. J. (2005). A positive behavioural intervention for toddlers: Parent–child attunement therapy. Child Abuse Review: Journal of the British Association for the Study and Prevention of Child Abuse and Neglect, 14(2), 132–151.

Dozier, M., Peloso, E., Lindhiem, O., Gordon, M. K., Manni, M., Sepulveda, S., … Levine, S. (2006). Developing evidence-based interventions for foster children: An example of a randomized clinical trial with infants and toddlers. Journal of Social Issues, 62(4), 767–785.

Dozier, M., Roben, C. K., Caron, E., Hoye, J., & Bernard, K. (2018). Attachment and biobehavioral catch-up: An evidence-based intervention for vulnerable infants and their families. Psychotherapy Research, 28(1), 18–29.

Evans, S., Davies, S., Williams, M., & Hutchings, J. (2015). Short-term benefits from the incredible years parents and babies programme in Powys. Community Practitioner, 88(9), 46.

Eyberg, S. M., Boggs, S. R., & Algina, J. (1995). Parent-child interaction therapy: a psychosocial model for the treatment of young children with conduct problem behavior and their families. Psychopharmacology bulletin.

Foreman, D. (2015). The psychiatry of children aged 0–4: advances in assessment, diagnosis and treatment. BJPsych Advances, 21(6), 377–386.

Foster, E. M., Prinz, R. J., Sanders, M. R., & Shapiro, C. J. (2008). The costs of a public health infrastructure for delivering parenting and family support. Children and Youth Services Review, 30(5), 493–501.

Fox, R., & Gresl, B. (2014). Early pathways: Home-based mental health services for young children in poverty. Unpublished training manual, Marquette University, Milwaukee, WI.

Fox, R. A., & Holtz, C. A. (2009). Treatment outcomes for toddlers with behaviour problems from families in poverty. Child and Adolescent Mental Health, 14(4), 183–189.

Fox, R. A., & Nicholson, B. (2003). Parenting young children: A facilitator’s guide.

French, A.N., Yates, B.T., & Fowles, T.R. (2018). Cost-Effectiveness of Parent–Child Interaction Therapy in Clinics versus Homes: Client, Provider, Administrator, and Overall Perspectives. Journal of Child and Family Studies, 27(10), 3329–3344.

Fung, M. P., & Fox, R. A. (2014). The culturally-adapted Early Pathways program for young Latino children in poverty: A randomized controlled trial. Journal of Latina/o Psychology, 2(3), 131.

Fusaro, V. A., Levy, H. G., & Shaefer, H. L. (2018). Racial and ethnic disparities in the lifetime prevalence of homelessness in the United States. Demography, 55(6), 2119–2128.

George, E., & Engel, L. (1980). The clinical application of the biopsychosocial model. American Journal of Psychiatry, 137(5), 535–544.

Gewirtz, A. H., DeGarmo, D. S., Plowman, E. J., August, G., & Realmuto, G. (2009). Parenting, parental mental health, and child functioning in families residing in supportive housing. American Journal of Orthopsychiatry, 79(3), 336–347.

Ghanbari, S., Khodapanahi, M., Gholamali Lavasani, M., Mazaheri, M., & Rezapour Faridian, R. (2018). The Effectiveness of Attachment-based Parent Training Method in Anxiety Syndrome of Preschool Children. International Journal of Behavioral Sciences, 12(1), 9–17.

Girard, E. I., Wallace, N. M., Kohlhoff, J. R., Morgan, S. S., & McNeil, C. B. (2018). Parent-Child Interaction Therapy with Toddlers.

Glasgow, R. E., Vogt, T. M., & Boles, S. M. (1999). Evaluating the public health impact of health promotion interventions: the RE-AIM framework. American journal of public health, 89(9), 1322–1327.

Glazemakers, I., & Deboutte, D. (2013). Modifying the ‘Positive Parenting Program’for parents with intellectual disabilities. Journal of Intellectual Disability Research, 57(7), 616–626.

Graziano, P. A., Ros-Demarize, R., & Hare, M. M. (2020). Condensing parent training: A randomized trial comparing the efficacy of a briefer, more intensive version of Parent-Child Interaction Therapy (I-PCIT). Journal of consulting and clinical psychology, 88(7), 669.

Grube, W. A., & Liming, K. W. (2018). Attachment and Biobehavioral Catch-up: A systematic review. Infant Mental Health Journal, 39(6), 656–673.

Guralnick, M. J. (2011). Why early intervention works: A systems perspective. Infants and young children, 24(1), 6.

Hare, M. M., & Graziano, P. A. (2020). The Cost-Effectiveness of Parent–Child Interaction Therapy: Examining Standard, Intensive, and Group Adaptations. Administration and Policy in Mental Health and Mental Health Services Research, 1–15.

Harris, S. E., Fox, R. A., & Love, J. R. (2015). Early pathways therapy for young children in poverty: A randomized controlled trial. Counseling Outcome Research and Evaluation, 6(1), 3–17.

Haskett, M. E., Armstrong, J., Neal, S. C., & Aldianto, K. (2018). Perceptions of triple P-positive parenting program seminars among parents experiencing homelessness. Journal of Child and Family Studies, 27(6), 1957–1967.

Herbers, J. E., Cutuli, J., Monn, A. R., Narayan, A. J., & Masten, A. S. (2014). Trauma, adversity, and parent–child relationships among young children experiencing homelessness. Journal of abnormal child psychology, 42(7), 1167–1174.

Homelessness, N. A. t. E. (2020). State of Homelessness: 2020 Edition. Retrieved from https://endhomelessness.org/homelessness-in-america/homelessness-statistics/state-of-homelessness-2020/

Hood, K. K., & Eyberg, S. M. (2003). Outcomes of parent-child interaction therapy: Mothers’ reports of maintenance three to six years after treatment. Journal of Clinical Child and Adolescent Psychology, 32(3), 419–429.

Ippen, C. G., Harris, W. W., Van Horn, P., & Lieberman, A. F. (2011). Traumatic and stressful events in early childhood: Can treatment help those at highest risk? Child abuse & neglect, 35(7), 504–513.

Jones, C. H., Erjavec, M., Viktor, S., & Hutchings, J. (2016). Outcomes of a comparison study into a group-based infant parenting programme. Journal of Child and Family Studies, 25(11), 3309–3321.

Juffer, F., BAKERMANS-KRANENBURG, M. J., & Van Ijzendoorn, M. H. (2018). Video-fedback intervention to promote positive parenting and sensitive discipline. Handbook of attachment-based interventions. New York: Guilford.

Juffer, F. E., Bakermans-Kranenburg, M. J., & Van Ijzendoorn, M. H. (2008). Promoting positive parenting: An attachment-based intervention: Taylor & Francis Group/Lawrence Erlbaum Associates.

Keeshin, B. R., Oxman, A., Schindler, S., & Campbell, K. A. (2015). A domestic violence shelter parent training program for mothers with young children. Journal of family violence, 30(4), 461–466.

Klein Velderman, M., Bakermans-Kranenburg, M. J., Juffer, F., Van Ijzendoorn, M. H., Mangelsdorf, S. C., & Zevalkink, J. (2006). Preventing preschool externalizing behavior problems through video-feedback intervention in infancy. Infant Mental Health Journal: Official Publication of The World Association for Infant Mental Health, 27(5), 466–493.

Knudsen, E. I. (2004). Sensitive periods in the development of the brain and behavior. Journal of cognitive neuroscience, 16(8), 1412–1425.

Kohlhoff, J., Morgan, S., Briggs, N., Egan, R., & Niec, L. (2020). Parent–Child Interaction Therapy with Toddlers: A Community-based Randomized Controlled Trial with Children Aged 14-24 Months. Journal of Clinical Child & Adolescent Psychology, 1–16.

Kushel, M. (2015). The first step is the hardest: overcoming barriers to primary care. In: Springer.

Leeb, R. T., Lewis, T., & Zolotor, A. J. (2011). A review of physical and mental health consequences of child abuse and neglect and implications for practice. American Journal of Lifestyle Medicine, 5(5), 454–468.

Leijten, P., Gardner, F., Landau, S., Harris, V., Mann, J., Hutchings, J., … Scott, S. (2018). Research Review: Harnessing the power of individual participant data in a meta-analysis of the benefits and harms of the Incredible Years parenting program. Journal of Child Psychology and Psychiatry, 59(2), 99–109.

Lieberman, A. F., Ippen, C. G., & Van Horn, P. (2006). Child-parent psychotherapy: 6-month follow-up of a randomized controlled trial. Journal of the American Academy of Child & Adolescent Psychiatry, 45(8), 913–918.

Lieberman, A. F., & Osofsky, J. D. (2009). Poverty, trauma, and infant mental health. Zero to Three, 30(2), 54.

Lieberman, A. F., Van Horn, P., & Ippen, C. G. (2005). Toward evidence-based treatment: Child-parent psychotherapy with preschoolers exposed to marital violence. Journal of the American Academy of Child & Adolescent Psychiatry, 44(12), 1241–1248.

Lieneman, C. C., Brabson, L. A., Highlander, A., Wallace, N. M., & McNeil, C. B. (2017). Parent–Child Interaction Therapy: Current Perspectives. Psychology Research and Behavior Management.

Lind, T., Bernard, K., Ross, E., & Dozier, M. (2014). Intervention effects on negative affect of CPS-referred children: Results of a randomized clinical trial. Child abuse & neglect, 38(9), 1459–1467.

Lind, T., Raby, K. L., Caron, E., Roben, C. K., & Dozier, M. (2017). Enhancing executive functioning among toddlers in foster care with an attachment-based intervention. Development and psychopathology, 29(2), 575.

Lippert, A. M., & Lee, B. A. (2015). Stress, coping, and mental health differences among homeless people. Sociological Inquiry, 85(3), 343–374.

Love, J. R., & Fox, R. A. (2019). Home-based parent child therapy for young traumatized children living in poverty: A randomized controlled trial. Journal of Child & Adolescent Trauma, 12(1), 73–83.

Lyons-Ruth, K., Todd Manly, J., Von Klitzing, K., Tamminen, T., Emde, R., Fitzgerald, H., … Foley, M. (2017). The worldwide burden of infant mental and emotional disorder: report of the task force of the World Association for infant mental health. Infant Mental Health Journal, 38(6), 695–705.

Madigan, S., Moran, G., Schuengel, C., Pederson, D. R., & Otten, R. (2007). Unresolved maternal attachment representations, disrupted maternal behavior and disorganized attachment in infancy: Links to toddler behavior problems. Journal of Child Psychology and Psychiatry, 48(10), 1042–1050.

Menting, A. T., de Castro, B. O., & Matthys, W. (2013). Effectiveness of the Incredible Years parent training to modify disruptive and prosocial child behavior: A meta-analytic review. Clinical Psychology Review, 33(8), 901–913.

Morpeth, L., Blower, S., Tobin, K., Taylor, R. S., Bywater, T., Edwards, R. T., … Berry, V. (2017). The effectiveness of the Incredible Years pre-school parenting programme in the United Kingdom: a pragmatic randomised controlled trial. Child Care in Practice, 23(2), 141–161.

Moss, E., Dubois-Comtois, K., Cyr, C., Tarabulsy, G. M., St-Laurent, D., & Bernier, A. (2011). Efficacy of a home-visiting intervention aimed at improving maternal sensitivity, child attachment, and behavioral outcomes for maltreated children: A randomized control trial. Development and psychopathology, 23(1), 195–210.

Nicholson, B., Anderson, M., Fox, R., & Brenner, V. (2002). One family at a time: A prevention program for at-risk parents. Journal of Counseling & Development, 80(3), 362–371.

Nowak, C., & Heinrichs, N. (2008). A comprehensive meta-analysis of Triple P-Positive Parenting Program using hierarchical linear modeling: Effectiveness and moderating variables. Clinical child and family psychology review, 11(3), 114.

Olds, D. L. (2006). The nurse–family partnership: An evidence-based preventive intervention. Infant Mental Health Journal, 27(1), 5–25.

Organization, W. H. (2004). Prevention of mental disorders: Effective interventions and policy options: Summary report: World Health Organization.

Park, J. M., Fertig, A. R., & Allison, P. D. (2011). Physical and mental health, cognitive development, and health care use by housing status of low-income young children in 20 American cities: A prospective cohort study. American journal of public health, 101(S1), S255–S261.

Peacock, S., Konrad, S., Watson, E., Nickel, D., & Muhajarine, N. (2013). Effectiveness of home visiting programs on child outcomes: a systematic review. BMC public health, 13(1), 17.

Pearl, E., Thieken, L., Olafson, E., Boat, B., Connelly, L., Barnes, J., & Putnam, F. (2012). Effectiveness of community dissemination of parent–child interaction therapy. Psychological Trauma: Theory, Research, Practice, and Policy, 4(2), 204.

Pearl, E. S. (2009). Parent management training for reducing oppositional and aggressive behavior in preschoolers. Aggression and Violent Behavior, 14(5), 295–305.

Prinz, R. J., Sanders, M. R., Shapiro, C. J., Whitaker, D. J., & Lutzker, J. R. (2009). Population-based prevention of child maltreatment: The US Triple P system population trial. Prevention Science, 10(1), 1–12.

Sampaio, F., Barendregt, J. J., Feldman, I., Lee, Y. Y., Sawyer, M. G., Dadds, M. R., … Mihalopoulos, C. (2018). Population cost-effectiveness of the Triple P parenting programme for the treatment of conduct disorder: an economic modelling study. European child & adolescent psychiatry, 27(7), 933–944.

Sampers, J., Anderson, K. G., Hartung, C. M., & Scambler, D. J. (2001). Parent training programs for young children with behavior problems. Infant-Toddler Intervention.

Sanders, M. R. (1999). Triple P-Positive Parenting Program: Towards an empirically validated multilevel parenting and family support strategy for the prevention of behavior and emotional problems in children. Clinical child and family psychology review, 2(2), 71–90.

Sanders, M. R., Ralph, A., Sofronoff, K., Gardiner, P., Thompson, R., Dwyer, S., & Bidwell, K. (2008). Every family: A population approach to reducing behavioral and emotional problems in children making the transition to school. The Journal of Primary Prevention, 29(3), 197–222.

Schoemaker, N. K., Juffer, F., Rippe, R. C., Vermeer, H. J., Stoltenborgh, M., Jagersma, G. J., … Alink, L. R. (2020). Positive parenting in foster care: Testing the effectiveness of a video-feedback intervention program on foster parents’ behavior and attitudes. Children and Youth Services Review, 110, 104779.

Seifer, R., & Dickstein, S. (2000). Parental mental illness and infant development.

Skovgaard, A. M., Houmann, T., Christiansen, E., Landorph, S., Jørgensen, T., Team, C. S., … Samberg, V. (2007). The prevalence of mental health problems in children 1½ years of age–the Copenhagen Child Cohort 2000. Journal of Child Psychology and Psychiatry, 48(1), 62–70.

Smith, M. (2004). Parental mental health: disruptions to parenting and outcomes for children. Child & Family Social Work, 9(1), 3–11.

Stronach, E. P., Toth, S. L., Rogosch, F., & Cicchetti, D. (2013). Preventive interventions and sustained attachment security in maltreated children. Development and psychopathology, 25(4pt1), 919–930.

Substance Abuse and Mental Health Services Administration (2020).Housing and Shelter.

Retrieved from https://www.samhsa.gov/homelessness-programs-resources/hpr-resources/housing-shelter

Sweet, M. A., & Appelbaum, M. I. (2004). Is home visiting an effective strategy? A meta-analytic review of home visiting programs for families with young children. Child development, 75(5), 1435–1456.

Szaniecki, E., & Barnes, J. (2016). Measurement issues: Measures of infant mental health. Child and Adolescent Mental Health, 21(1), 64–74.

Tabachnick, A. R., Raby, K. L., Goldstein, A., Zajac, L., & Dozier, M. (2019). Effects of an attachment-based intervention in infancy on children’s autonomic regulation during middle childhood. Biological psychology, 143, 22–31.

Thomas, R., Abell, B., Webb, H. J., Avdagic, E., & Zimmer-Gembeck, M. J. (2017). Parent-child interaction therapy: A meta-analysis. Pediatrics, 140(3), e20170352.

Thomas, R., & Zimmer-Gembeck, M. J. (2012). Parent–Child Interaction Therapy: An evidence-based treatment for child maltreatment. Child maltreatment, 17(3), 253–266.

Tischler, V., Rademeyer, A., & Vostanis, P. (2007). Mothers experiencing homelessness: Mental health, support and social care needs. Health & social care in the community, 15(3), 246–253.

Toomey, E., Matvienko-Sikar, K., Heary, C., Delaney, L., Queally, M., Hayes, C. B., … xteam, C. H. E. f. I. H. s. (2019). Intervention fidelity within trials of infant feeding behavioral interventions to prevent childhood obesity: a systematic review. Annals of Behavioral Medicine, 53(1), 75–97.

Toth, S. L., & Gravener, J. (2012). Bridging research and practice: Relational interventions for maltreated children. Child and Adolescent Mental Health, 17(3), 131–138.

Toth, S. L., Sturge-Apple, M. L., Rogosch, F. A., & Cicchetti, D. (2015). Mechanisms of change: Testing how preventative interventions impact psychological and physiological stress functioning in mothers in neglectful families. Development and psychopathology, 27(4 0 2), 1661.

Tully, L. A., & Hunt, C. (2016). Brief parenting interventions for children at risk of externalizing behavior problems: A systematic review. Journal of Child and Family Studies, 25(3), 705–719.

U.S. Department of Housing and Urban Development. (2012). A Guide to Counting Sheltered Homeless People. Retrieved from https://www.hudexchange.info/sites/onecpd/assets/File/A-Guide-to-Counting-Sheltered.pdf

Uylings, H. B. (2006). Development of the human cortex and the concept of “critical” or “sensitive” periods. Language Learning, 56, 59–90.

van der Asdonk, S., Cyr, C., & Alink, L. (2020). Improving parent–child interactions in maltreating families with the Attachment Video-feedback Intervention: Parental childhood trauma as a moderator of treatment effects. Attachment & Human Development, 1–21.

van IJzendoorn, M. H., Juffer, F., & Duyvesteyn, M. G. (1995). Breaking the intergenerational cycle of insecure attachment: A review of the effects of attachment-based interventions on maternal sensitivity and infant security. Journal of Child Psychology and Psychiatry, 36(2), 225–248.

Van Zeijl, J., Mesman, J., Stolk, M. N., Alink, L. R., Van IJzendoorn, M. H., Bakermans-Kranenburg, M. J., … Koot, H. M. (2006). Terrible ones? Assessment of externalizing behaviors in infancy with the Child Behavior Checklist. Journal of Child Psychology and Psychiatry, 47(8), 801–810.

Ward, M. A., Theule, J., & Cheung, K. (2016). Parent–child interaction therapy for child disruptive behaviour disorders: A meta-analysis. Paper presented at the Child & Youth Care Forum.

Webster-Stratton, Jamila Reid, M., & Stoolmiller, M. (2008). Preventing conduct problems and improving school readiness: evaluation of the Incredible Years Teacher and Child Training Programs in high-risk schools. Journal of child psychology and psychiatry, and allied disciplines, 49(5), 471–488. doi:10.1111/j.1469-7610.2007.01861.x

Webster-Stratton, C. (2017). Is the Incredible Years a trauma-focused therapy? Can IY parent and child programs be used for families where children have experienced trauma. The Incredible Years.

Webster-Stratton, C., & Reid, M. J. (2018). The Incredible Years parents, teachers, and children training series: A multifaceted treatment approach for young children with conduct problems.

Webster-Stratton, C., Jamila Reid, M., & Stoolmiller, M. (2008). Preventing conduct problems and improving school readiness: evaluation of the incredible years teacher and child training programs in high-risk schools. Journal of Child Psychology and Psychiatry, 49(5), 471–488.

Weiner, D. A., Schneider, A., & Lyons, J. S. (2009). Evidence-based treatments for trauma among culturally diverse foster care youth: Treatment retention and outcomes. Children and Youth Services Review, 31(11), 1199–1205.

Weisz, J. R., Sandler, I. N., Durlak, J. A., & Anton, B. S. (2005). Promoting and protecting youth mental health through evidence-based prevention and treatment. American psychologist, 60(6), 628.

Wessels, I., & Ward, C. L. (2016). Battered women and parenting: acceptability of an evidence-based parenting programme to women in shelters. Journal of Child & Adolescent Mental Health, 28(1), 21–31.

Wichstrøm, L., Berg-Nielsen, T. S., Angold, A., Egger, H. L., Solheim, E., & Sveen, T. H. (2012). Prevalence of psychiatric disorders in preschoolers. Journal of Child Psychology and Psychiatry, 53(6), 695–705.

Wiggins, T. L., Sofronoff, K., & Sanders, M. R. (2009). Pathways triple P-positive parenting program: effects on parent-child relationships and child behavior problems. Family Process, 48(4), 517–530.

Williams, M. E. (2016). Integrating early childhood mental health and trauma-informed care for homeless families with young children. Pragmatic Case Studies in Psychotherapy, 12(2), 113–123.

Wilson, P., Rush, R., Hussey, S., Puckering, C., Sim, F., Allely, C. S., … Gillberg, C. (2012). How evidence-based is an’evidence-based parenting program’? A PRISMA systematic review and meta-analysis of Triple P. BMC medicine, 10(1), 130.

Wood, D. L., Valdez, R. B., Hayashi, T., & Shen, A. (1990). Health of homeless children and housed, poor children. Pediatrics, 86(6), 858–866.

Zima, B. T., Wells, K. B., Benjamin, B., & Duan, N. (1996). Mental health problems among homeless mothers: Relationship to service use and child mental health problems. Archives of General Psychiatry, 53(4), 332–338.

Zisser, A., & Eyberg, S. M. (2010). Parent-child interaction therapy and the treatment of disruptive behavior disorders.

